# Multi-omics of sorafenib responsiveness in HCC patients

**DOI:** 10.1101/2025.07.15.25331324

**Authors:** Eva Dazert, Tujana Boldanova, Charlotte KY Ng, Marco Colombi, Stefan Wieland, George Rosenberger, Francesco Marass, Salvatore Piscuoglio, Niko Beerenwinkel, Luigi Terracciano, Markus Heim, Michael N. Hall

**Affiliations:** Biozentrum, University of Basel; Basel, Switzerland; Department of Biomedicine, University Hospital Basel; Basel, Switzerland; Department of Gastroenterology and Hepatology, University Hospital Basel; Basel, Switzerland; Department for BioMedical Research (DBMR), University of Bern; Bern, Switzerland; Institute of Molecular Genetics and Pathology, University Hospital Basel; Basel, Switzerland; SIB Swiss Institute of Bioinformatics; Lausanne, Switzerland; Department of Biosystems Science and Engineering, ETH Zürich; Basel, Switzerland; SIB Swiss Institute of Bioinformatics; Basel, Switzerland; Department of Pathology, Humanitas Clinical and Research Center, IRCCS; Milan, Italy; Humanitas University, Department of Biomedical Sciences; Milan, Italy

**Author notes:** The authors declare no financial conflict of interest.

## Abstract

Tumor response rates to targeted therapy are generally low. This poses an urgent medical need to identify molecular mechanisms determining responsiveness to inform therapy. We describe the first longitudinal deepscale multi-omic analysis (genomics, transcriptomics, proteomics and phosphoproteomics) of tumor biopsies from eight hepatocellular carcinoma (HCC) patients treated with the targeted kinase inhibitor sorafenib. Three patients were sorafenib responders and five were nonresponders. Resistance did not correlate with a single mutated gene but with genomic instability, especially gain of Chr1q. The patients clustered by therapy response, based on proteomics and phosphoproteomics. Nonresponder tumors showed increased EMT, glycogen storage, splicing and ribosome biogenesis, and decreased carbohydrate and drug metabolism. Resistance also correlated with deregulated phosphorylation of AMPK and NOTCH pathway components. Our data suggest a correlation between dedifferentiation and resistance not evident based on classical clinical staging. We propose potential novel biomarkers to identify and drug combinations to treat sorafenib resistant patients.

**STATEMENT OF SIGNIFICANCE:** We provide potential mechanisms of responsiveness to cancer therapy by deep-scale multi-omic analysis of longitudinal tumor biopsies from eight hepatocellular carcinoma (HCC) patients treated with sorafenib.

## INTRODUCTION

Hepatocellular carcinoma (HCC) as the most common form of liver cancer, is the fourth-leading cause of cancer-related death and a global health concern (Villanueva, 2019). The 5-year survival rate of liver cancer is only 18%. The prognosis for HCC patients is poor with only 30% qualifying for curative treatments such as tumor resection or transplantation (Gallage *et al*, 2021; Mazzanti *et al*, 2008). Loco-regional therapies using radiofrequency or chemoablation, are available to patients with preserved liver functions not suitable for surgical intervention. Most HCC patients are diagnosed at advanced stage, where only systemic therapies are recommended. Mutations driving HCC affect telomere maintenance (TERT), cell cycle control (TP53), Wnt/beta-catenin signaling (CTNNB1), oxidative stress pathway (NFE2L2), epigenetic modifiers (ARID1A) and proliferative pathways (mTOR/MAPK) (Zucman-Rossi *et al*, 2015). However, clinical translation of this information is hampered by lack of mutation-targeting drugs and poor characterization of indirect mutational effects.

Sorafenib, the first first-line targeted therapy for HCC, is a multi-kinase inhibitor (MKI) targeting B-Raf, C-Raf, VEGFR, PDGFR, RET and KIT (Wilhelm *et al*, 2004). It prolongs median overall survival (OS) and progression free survival (PFS) on average by 3 months (10.7 vs 7.9 months with placebo) and was introduced in 2008 (SHARP trial) (Llovet *et al*, 2008). Lenvatinib as MKI with similar targets as sorafenib, was introduced in 2018 as first/second-line therapy for HCC due to its improvement of PFS by approx. 5 months (9 vs 4 month with sorafenib) (Kudo *et al*, 2018). In 2020, the IMBRAVE-150 trial combining atezolizumab and bevacizumab (immunotherapy) showed an increase of median OS by approx. 6 months (19 vs 13 month with sorafenib) (Finn *et al*, 2021; Finn *et al*, 2020), recommending it as novel first-line therapy for HCC. The treatment landscape of HCC has therefore changed largely in the very recent past. Sorafenib is currently used as second-line therapy. However, clinical benefit of the systemic HCC therapy options is still limited, due to lack of drugs targeting pathways deregulated in HCC and to intrinsic or acquired resistance to therapy. Interestingly, although most patients die within few months after start of targeted therapy, e.g. with sorafenib, few patients respond and survive for years under sorafenib. Predictive biomarkers to stratify HCC patients for targeted therapy are scarce and not present for HCC. The identification of mechanisms underlying responsiveness to systemic targeted therapy based on kinase inhibition are therefore of high medical need. Here, we aim at unraveling such mechanisms by analyzing sorafenib responder and nonresponder HCC patients. Such insights would not only be of general importance to understand mechanisms of responsiveness to targeted therapy, but would also aid in second-line upfront identification of sorafenib nonresponders to prevent toxicity of ineffective therapy. Previous studies in model systems and patients suggest that resistance to targeted therapy is mediated by efflux pumps, alternative proliferative pathways, metabolic symbiosis, autophagy or EMT (van Malenstein *et al*, 2013b; Zhai *et al*, 2014). Sorafenib resistance correlates with upregulation of the mTOR pathway (Masuda *et al*, 2014) or MAPK14 (Rudalska *et al*, 2014). Longitudinal biopsy studies in melanoma showed that resistance to B-Raf inhibition occurs via mutational activation of the B-Raf effector kinase MEK (Wagle *et al*, 2011).

Attempts to classify HCC into molecular subgroups to guide treatment have had limited success, possibly because they have relied on resected tumor material available only from early-stage HCC patients (Gao *et al*, 2019; Jiang *et al*, 2019) or are often based solely on transcriptomic data (Boyault *et al*, 2007; Goossens *et al*, 2015; Hoshida *et al*, 2009; Lee *et al*, 2004). Based on these studies, the current notion divides HCC in two main classes, proliferative and nonproliferative, which have been further subdivided in 6 groups based on further more specific molecular differences between the groups (Gallage *et al*., 2021). Landmark studies have combined genomics and proteomics to examine resected HCC (Gao *et al*., 2019; Jiang *et al*., 2019) and other solid cancer types (Clark *et al*, 2019; Gillette *et al*, 2020; McDermott *et al*, 2020) and, in a proof-of-concept study, breast cancer biopsies taken pre- and 72 hours post-treatment (Satpathy *et al*, 2020). However, a multi-omic study that combines genomics, transcriptomics, proteomics and phosphoproteomics (p-proteomics) to examine longitudinal HCC biopsies from sorafenib sensitive and resistant patients has not been performed.

To unravel molecular mechanisms underlying responsiveness to sorafenib in vivo, we performed a longitudinal multi-omic study on a cohort of eight sorafenib-treated HCC patients. We analyzed tumor (TU) and matched nontumor (NT) biopsies obtained before and during treatment from three responder and five nonresponder patients. We found that the proteomes and p-proteomes of the patients clustered by therapy response, and identified several signaling pathways as potential contributors to resistance. We observed a correlation of sorafenib resistance and dedifferentiation not evident by classical clinical staging.

## RESULTS

### Longitudinal cohort of eight sorafenib-treated HCC patients

To uncover molecular determinants of responsiveness to sorafenib, we analyzed eight sorafenib-treated HCC patients (Figure 1SA and Table S1), in particular three responders and five nonresponders. According to our nomenclature convention, R and NR refer to the responder and nonresponder groups, respectively, whereas NR1, NR2 etc. refers to a specific patient from the corresponding group. We did not find a clinical covariate predicting responsiveness to sorafenib therapy, except the expected significant difference in survival time post treatment start (p=0.04817321, log-rank test). On average, R survived 100 weeks longer than NR (Figure S1B). A detailed overview of the clinical parameters of all patients and biopsies analyzed in this study can be seen in Table S1. Each patient sample set consisted of at least four biopsies, one pre- and one ontreatment biopsy pair taken from the TU and matched NT tissue. Four patient sample sets (R1, R3, NR3, and NR5) included more than four biopsies, corresponding to additional time points or tumors (e.g., portal vein metastases). We performed multi-omic analysis of the TU and NT biopsies, including genomics, transcriptomics, proteomics and p-proteomics (Figure S1C). Biopsies were immediately snap-frozen upon collection to preserve the in vivo status of normally labile p-proteomes.

**Fig. 1:**
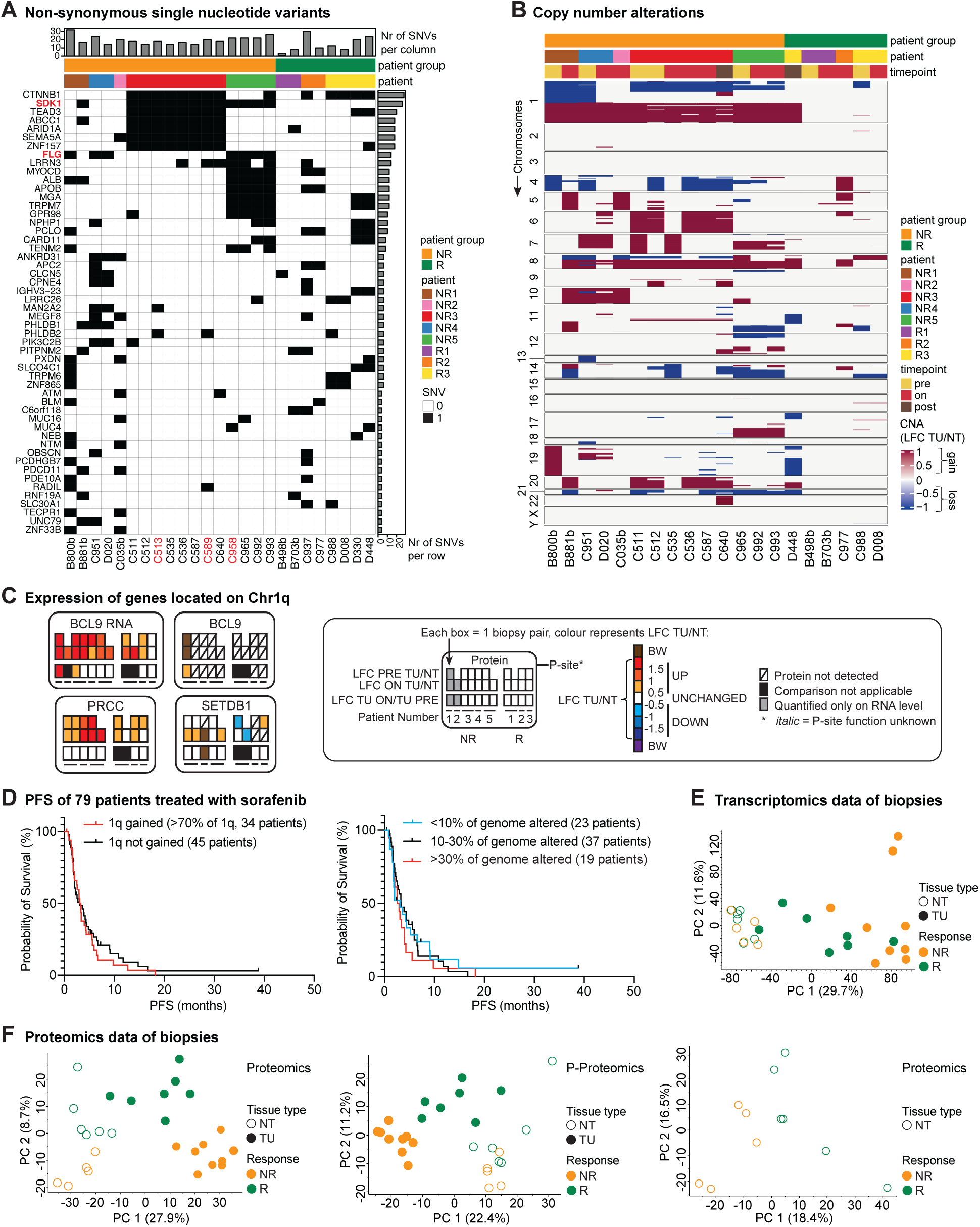
Proteomics reveal differences between responder and nonresponder patients at greater depth than genomics and transcriptomics. **(A)** Non-synonymous SNVs present in the TU biopsies of a minimum of two patients. Portal vein metastases biopsies are labeled in red. **(B)** CNAs ordered by therapy response and location on chromosomes (left side from top to bottom). Gains = red, losses = blue. **(C)** Omics tiles for concordant genes BCL9, PRCC and SETDB1 which were also present in COSMIC database. Each box displays the calculated LFC data from one biopsy pair, namely pretreatment TU/NT in the first row, ontreatment TU/NT in the second row and TU ontreatment/TU pretreatment in the third row. For NR2, only the ontreatment biopsy pair could be analyzed. For R1, more biopsies could be collected at the ontreatment time point then the pretreatment time point, therefore one position is empty in the first row. From NR1 and NR2 only transcriptomics data are available, hence the tile is named RNA if the transcriptome data are displayed. See legend in box for more details. **(D)** Kaplan-Meier curve for PFS of sorafenib-treated patients from an independent cohort (Harding *et al*., 2019). Left panel shows comparison of patients with or without gain in Chr1q. Right panel shows comparison of three groups of patients harbouring different degree of genomic instability. **(E)** PCA of transcriptomics data from main biopsies (TU and NT biopsies). Shown are mean ratios of each biopsy i.e., mean of triplicate measurements of the normalized ratio H/L. **(F)** PCA of proteomics data from main biopsies. Left panel: Mean ratios of proteomics data of TU and NT biopsies. Middle panel: Mean ratios of p-proteomics data of TU and NT biopsies. Right panel: Mean ratios of proteomics data of NT biopsies alone. Shown are mean ratios of each biopsy i.e., mean of triplicate measurements of the normalized ratio H/L.

### Responsiveness to targeted therapy cannot be explained by a single gene

We found single nucleotide variants (SNVs) (Fig. S1D) in genes commonly mutated in HCC, e.g., *CTNNB1* and *ARID1A*, in each case in both R and NR, suggesting that such genes do not determine responsiveness. Next, we extracted SNVs appearing exclusively in R or NR, in each case in at least three patients. *FLG* and *SDK1* (Fig. 1A) contained non-synonymous SNVs in three of the five NR patients. In conclusion, although mutations in *FLG* and *SDK1* appeared only in NR and may thus contribute to sorafenib resistance, there is no gene that when mutated is absolutely required for resistance.

### Resistance correlates with genomic instability and gain of chromosome arm 1q

NR displayed a higher number of copy number alterations (CNAs) (approx. 5000) compared to R (approx. 1000) (Fig. S1E). Importantly, all NR tumors gained a complete chromosome arm 1q (Fig. 1B), while a subset of NR gained 8q or lost 1p. These CNAs, observed in 40-60% of HCCs (Franch-Exposito *et al*, 2020), suggest increased genomic instability in NR. Genetic instability has been linked to sorafenib resistance (Ming-Chin Yu & Jang-Hau Lian, 2019) and poor prognosis in HCC. 23 genes on 1q are deemed cancer-relevant (COSMIC Cancer Gene Census). Of these, expression of *BCL9*, *PRCC* and *SETDB1* was upregulated in NR, based on log_2_ fold change (LFC) of TU vs NT (Fig. 1C). Next, we interrogated the genome sequences of an independent cohort of 127 HCC patients. 79 were treated with sorafenib of which 34 harboured a 1q gain (Harding *et al*, 2019). However, both 1q-gain and genome instability (>30%) conferred only a minor, not significant, decrease in progression-free survival (PFS) under sorafenib treatment compared to control patients (Fig. 1D). In conclusion, genomic instability alone, in particular 1q gain, may not be sufficient to predict sorafenib resistance.

### Patients cluster by therapy response, based on proteome and p-proteome but not transcriptome

Next, we examined our transcriptomic, proteomic and p-proteomic data, by principal component analysis (PCA) and hierarchical clustering (HC). Patients did not cluster by therapy response based on transcriptome (Fig. 1E and S2A) but did cluster based on proteome (Fig. 1F, S2B and S2D) and p-proteome (Fig. 1F, S2C and S2E). Unlike what is observed with therapy response (PC2), we note that biopsies segregated by tissue type (NT/TU) (PC1) in all omics data. Interestingly, NT biopsies also segregated by therapy response (PC2) (Fig. 1F), suggesting that nontumor properties contribute to responsiveness. In summary, only (p)proteomics capture molecular differences mediating responsiveness, independent of inter-patient variation.

### Nonresponder patients display increased EMT, ribosome biogenesis and Rho-GTPase signaling and decreased carbohydrate and drug metabolism

Next, we performed pathway enrichment analysis (PEA) of the proteome data to determine pathways differentially regulated between NR and R, pre- and/or on-treatment. Importantly, we normalized the tumor data from each biopsy with the matched non-tumor biopsy data to equalize for potential patient-specific expression changes and underlying tumor-independent but disease-related changes, for example cirrhosis etc. We then calculated the log2 fold change of TU vs NT biopsy for each p Up- and down-regulated proteins were analyzed separately and together. (Fig. 2A and 2B, Fig. S3A).

**Fig. 2:**
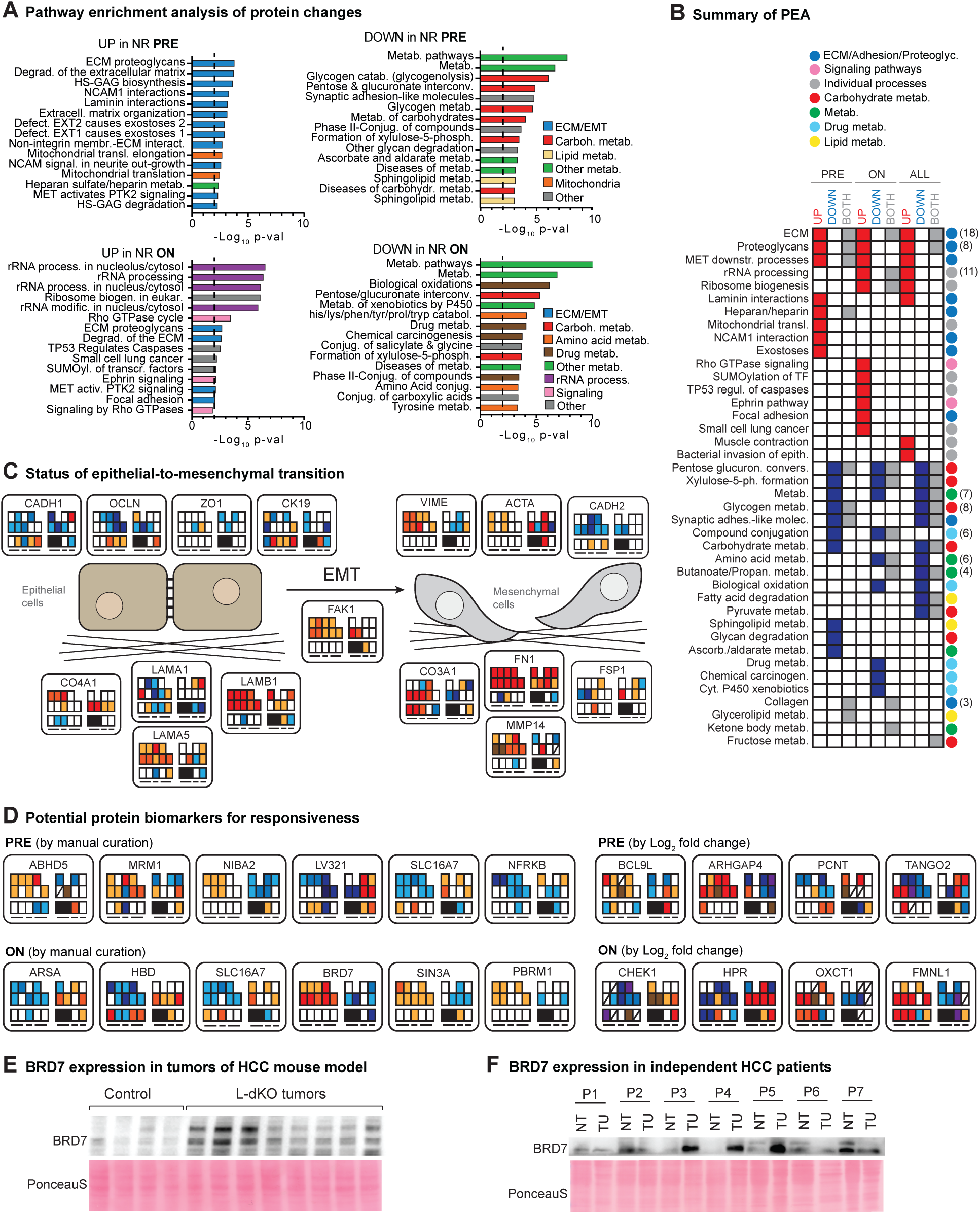
Nonresponder patients display increased EMT, ribosome biogenesis and Rho-GTPase signaling and decreased carbohydrate and drug metabolism. **(A)** Pathway enrichment analysis of protein changes in proteome data. Upper row: PEA of Top 500 deregulated proteins differentially expressed pretreatment between NR and R based on p-value and LFC of TU vs NT. Left panel: Upregulated proteins, right panel: downregulated proteins. Lower row: PEA of Top 500 deregulated proteins differentially expressed ontreatment between NR and R based on p-value and LFC of TU vs NT. Left panel: Upregulated proteins, right panel: downregulated proteins. Within timepoints, pathway annotations belonging to same cellular process were labeled with same colour. Dashed line indicates significance threshold. **(B)** Summary of PEA of proteome data pre-, ontreatment and overall (using all timepoints together), each time showing PEA results of upregulated (red), downregulated (blue) or up- and downregulated proteins together (grey). Enrichment pathways belonging to same term were added under one keyword (legend above heatmap), number of how many terms were added together is shown in brackets (right side of heatmap). Pathways belonging to same biological process were labeled as circles in the same colour (right side of heatmap). **(C)** Omics tiles for proteins involved in EMT which were quantified in this study cohort. See Figure 1C for legend of omics tiles. **(D)** Omics tiles of potential protein biomarkers for responsiveness. See Figure 1C for legend of omics tiles. Shown are strongest deregulated proteins between NR and R. Upper row: Pretreatment candidates extracted from Fig. S3A and S3B. Lower row: Ontreatment candidates extracted from Fig. S3D and S3E. Although candidates have been picked either from Top 50 list of the pre- or ontreatment analysis, all omics tiles of each protein are displayed. SLC16A7 is shown twice as it was a Top 50 hit in both the pre- and ontreatment analysis. Left side: candidates identified by manual curation of our Filemaker multi-omic database, right side: candidates identified by LFC search (for details see method section). **(E)** Immunoblot of BRD7 in L-dKO mouse tumors (HCC mouse model with liver-specific knockout of PTEN and TSC1) compared to control liver tissues (CTRL). Ponceau S image serves as loading control. N=4 (CTRL), N=4 (two tumors per L-dKO mouse). **(F)** Immunoblot of BRD7 in resected tumor (TU) tissue and adjacent nontumor (NT) tissue from seven human HCC patients. Ponceau S image serves as loading control.

Regarding pathways changed in NR both pre- and on-treatment (intrinsic resistance), extracellular matrix (ECM), cell adhesion and proteoglycan-related proteins were upregulated, while metabolic proteins, in particular involved in carbohydrate metabolism, were downregulated. The term ECM contains proteins involved in epithelial-to-mesenchymal transition (EMT) (Gonzalez & Medici, 2014). Regarding EMT (Fig. 2C), the epithelial markers CADH1, ZO1, OCLN, CK19 were downregulated pre- and on-treatment in both NR and R. Conversely, the mesenchymal markers VIME, FAK1, CO3A1 and MMP14 were upregulated, pre- and on-treatment, but only in NR. These data suggest that NR tumors had progressed further toward EMT compared to R tumors. EMT is associated with a cancer stem cell phenotype including resistance to targeted therapy (van Malenstein *et al*, 2013a). Proteins corresponding to carbohydrate, glycogen (glycogenolysis), glycan and sphingolipid metabolism, and compound conjugation were downregulated in NR pretreatment. Cancer cells accumulate glycogen as an energy source to survive under hypoxia and glucose deprivation (Zois & Harris, 2016). The downregulation of alternative glucose utilizing pathways, e.g., the Leloir pathway, suggests dependence on glycolysis in ontreatment NR tumors.

Regarding pathways changed in NR only ontreatment (acquired resistance), rRNA processing, ribosome biogenesis and Rho GTPase signaling were up- and drug metabolism was down-regulated. Ribosome biogenesis, possibly via ‘oncogenic’ ribosomes specialized for translation of cancer-relevant mRNAs (Bastide & David, 2018), might promote tumor growth. The term Rho GTPases contains RAC2 and ARHGEF1/2/7, suggesting stimulation of Rac GTPases. Regarding drug metabolism, it has been shown that the liver loses expression of several CYPs and UGTs involved in drug/xenobiotics metabolism upon dedifferentiation (Elaut *et al*, 2006). CYP3A4 and UGT1A9, the main metabolizers of sorafenib, are decreased in HCC(Ye *et al*, 2014) and also in our NR cohort at both time points. Collectively, the above suggests that EMT and adaptation to nutrient stress contribute to intrinsic resistance, while ribosome biogenesis, RhoGTPase signaling and reduced drug metabolism contribute to acquired resistance. The above also supports the notion that resistant tumors are more dedifferentiated.

Next, we searched for potential novel protein biomarkers for responsiveness (Fig. 2D, Fig. S3B-E and Fig.S4A-C). To this end, we performed two different searches for the Top 50 deregulated proteins between NR and R. In one search we filtered for the Top 50 deregulated candidate markers with the highest LFC of TU vs NT (e.g. pretreatment) in combination with the best p-value and from those extracted the candidates with the most extreme patterns of regulation (upregulated in NR and downregulated in R or vice versa). In the second search we filtered for the most extreme patterns of deregulation (upregulated in NR and downregulated in R or vice versa) by manual curation of our Filemaker multi-omics database. The hit criteria here were one of the following: 1) LFC>+/-0.75 and significant, or 2) LFC>+/-1 but not necessarily significant, or 3) black-and-white expression which means present in all triplicate measurements of TU and absent in all triplicate measurements of NT or vice versa (see method section). Of note, those candidates are suggested as potential biomarkers due to their differential expression in our study cohort but would need further validation in other cohorts. SLC16A7 was downregulated in NR and upregulated in R while SLC16A3 was inversely regulated. SLC16A7 and SLC16A3 correlate with good and bad prognosis, respectively (Alves *et al*, 2014). PCNT and SPC24, both downregulated in NR, are involved in chromosome segregation and possibly related to genome instability (see above). BCL9L, upregulated in NR, promotes CTNNB1 transcriptional activity to stimulate tumorigenesis (Huge *et al*, 2020). ARHGAP4, increased in NR, is a member of the rhoGAP family that stimulates Ras and thereby MAPK signaling. The transcriptional regulators BRD7, PBRM1 (both components of the PBAF SWI/SNF chromatin remodeling complex), ELF1 and SIN3A were upregulated in NR. From all the potential protein biomarkers for responsiveness, BRD7 showed the most clear and extreme deregulation between NR and R and has been described as a tumor suppressor. We therefore validated BRD7 expression in HCC. BRD7 expression was also upregulated in tumors of an mTOR-driven mouse model for aggressive HCC (Liver-specific PTEN/TSC1 deletion) compared to wildtype livers of floxed littermates (Fig. 2E). This result argues for a rather tumor promoting role of BRD7. We also tested BRD7 expression in resected tumor and adjacent non-tumor material from seven HCC patients. Interestingly, it was upregulated in three patients, unchanged in one patient and downregulated in three patients (Fig. 2F), therefore paralleling the expression pattern seen in our study cohort. In conclusion, these suggested biomarkers may be helpful for patient stratification and provide molecular insight on sorafenib responsiveness.

### Nonresponder patients show deregulated NOTCH signaling

Next, we asked which signaling pathways are differentially regulated in NR compared to R in our p-proteomic dataset.We therefore performed again PEA but now on differentially phosphorylated proteins, as revealed by our p-proteomic analysis (Fig. 3A and 3B, Fig. S4D). Regarding proteins whose phosphorylation was changed in NR both pre- and on-treatment, phosphorylation was increased in proteins involved in mRNA transport, processing and splicing. Conversely, phosphorylation was decreased in proteins associated with MAPK/mTOR signaling. Phosphorylation of proteins involved in NOTCH signaling was decreased pre-treatment and increased on-treatment. Finally, regarding proteins whose phosphorylation was changed only ontreatment, proteins involved in cell cycle/mitosis were upregulated and proteins involved in translation initiation were downregulated.

**Fig. 3:**
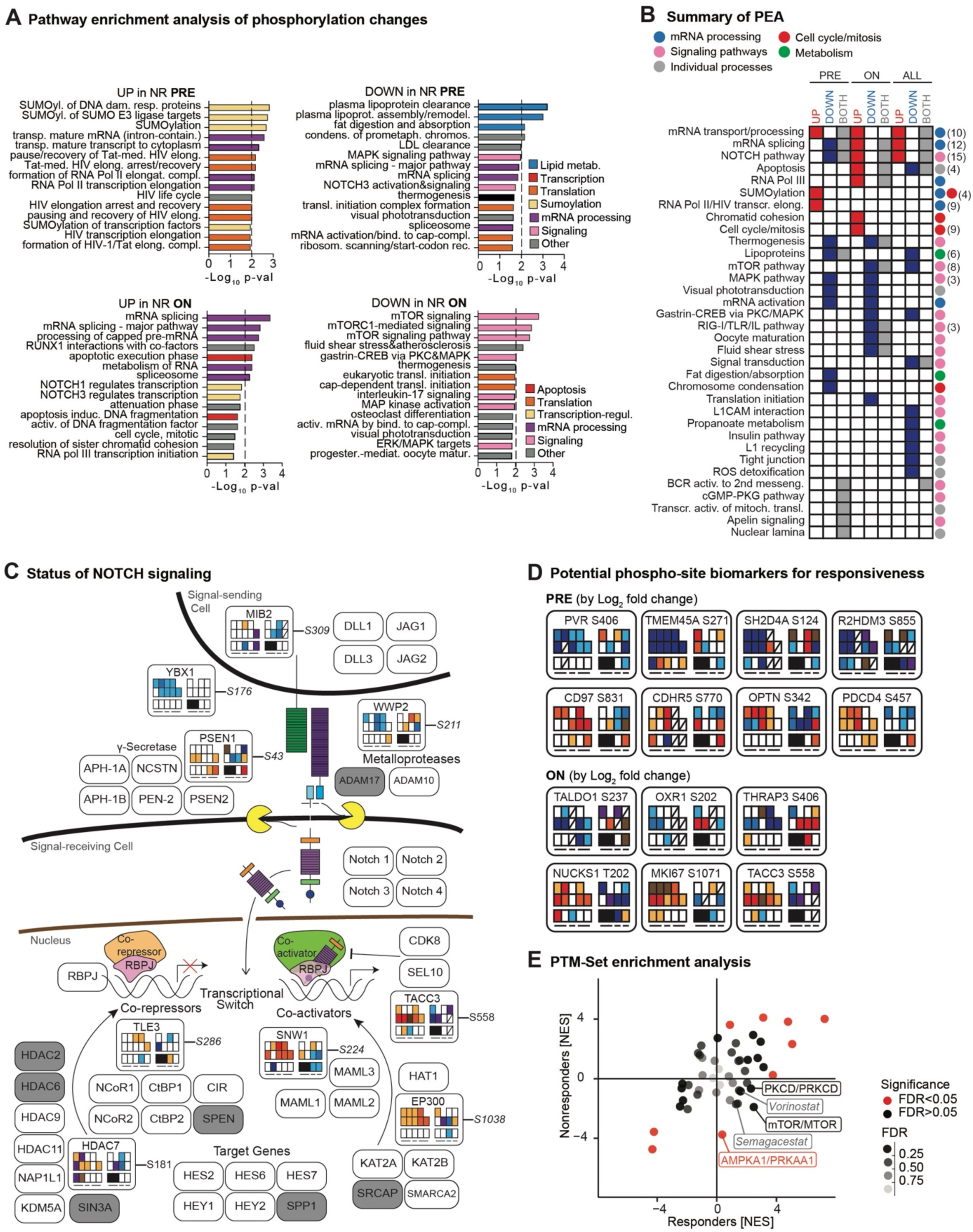
Nonresponder patients show deregulated NOTCH signaling and reduced AMPK signaling. **(A)** Pathway enrichment analysis of phosphorylation changes (taking into account phosphorylated proteins). Upper row: PEA of Top 300 phosphorylations deregulated pretreatment between NR and R based on p-value and LFC of TU vs NT. Left panel: Upregulated phosphorylations, right panel: downregulated phosphorylations. Lower row: PEA of Top 250 phosphorylations deregulated ontreatment between NR and R based on p-value and LFC of TU vs NT. Left panel: Upregulated phosphorylations, right panel: downregulated phosphorylations. Within timepoints, pathway annotations belonging to same cellular process were labeled with same colour. Dashed line indicates significance threshold. **(B)** Summary of PEA of p-proteome data pre-, ontreatment and overall (using all timepoints together), each time showing PEA results of upregulated (red), downregulated (blue) or up- and downregulated phosphorylations together (grey). Enrichment pathways belonging to same term were added under one keyword (legend above heatmap), number of how many terms were added together is shown in brackets (right side of heatmap). Pathways belonging to same biological process were labeled as circles in the same colour (right side of heatmap). **(C)** Omics tiles for p-sites of the NOTCH pathway deregulated in this study (adapted from (Borggrefe & Oswald, 2009; Bray, 2006, 2016; Kopan & Ilagan, 2009)). Only proteins with p-sites picked up by the PEA are depicted. White squares = NOTCH pathway components, grey squares= NOTCH components with p-sites detected in this study, but not picked up in PEA. **(D)** Omics tiles of potential p-site biomarkers for responsiveness. Shown are strongest deregulated phosphorylations between NR and R. Upper panel: Pretreatment candidates extracted from Fig. S4D and S4E, shown are candidates identified by LFC search. Lower panel: Ontreatment candidates extracted from Fig. S5A and S5B, shown are candidates identified by LFC search. **(E)** PTM-SEA analysis of p-sites deregulated between NR and R, taking into account the actual p-sites, not only the phosphorylated protein, here from all timepoints together. Enriched pathways are shown by their Normalized enrichment score (NES). Only points in discordant quadrants (upregulated in R and downregulated in NR) with an FDR<0.25 were labeled.

We next investigated the functional outcomes of the observed phosphorylation changes in their respective signaling pathway. We focused on those signaling pathways that appeared most often in the PEA, namely NOTCH, mTOR and MAPK signaling (Fig. 3B), since proteins belonging to them were overrepresented among the deregulated p-sites (see methods section). We retrieved the information about which p-sites belonged to which pathway from the PEA and visualized their quantitative data as omics tiles. The MAPK pathway (Fig. S5A) displayed a downregulation of early inhibitory p-sites in CRAF and BRAF in both patient groups. Activating p-sites in the central kinases MAPK3/1 were upregulated in NR and downregulated or undetected in R tumors. Phosphorylation of MAPK3/1 downstream target p-sites varied. Only phosphorylation of FMNA S2152, HMG14 S7 and CJUN S73 tended to be increased in NR. In general, sorafenib treatment only modestly affected MAPK signaling. Regarding the status of mTOR signaling (Fig. S5B), many inhibitory p-sites were downregulated, e.g, RPTOR S722, ACACA S80 and GSK3B S9, but the phosphorylation status of important mTOR-related targets (CAD, NDRG2, EIF4EBP1, RPS6) did not show a correlation with responsiveness in NR and R. Regarding the status of NOTCH signaling (Fig. 3D), NR showed decreased phosphorylation of YBX1 and WWP2 and increased phosphorylation of TACC3, EP300, SNW1, TLE3 and HDAC7 independent of treatment. Only phosphorylation of PSEN1 increased in NR ontreatment, arguing for an intrinsic deregulated NOTCH signaling in NR and R. From the three main deregulated signaling pathways in our p-proteomic data, NOTCH signaling components were most homogenously deregulated between NR and R. Collectively, we conclude that deregulated phosphorylation of proteins involved in mRNA transport/processing/splicing and non-canonical NOTCH signaling may contribute to intrinsic resistance, while proteins involved cell cycle/mitosis and translation initiation may contribute to acquired resistance in our study cohort.

Next, we searched for p-sites as potential biomarkers for responsiveness (Fig. 3C, Fig. S5C and Fig. S6A-D). Here, we performed again two searches as mentioned above but this time searched the deregulated p-sites as revealed by our p-proteomic data. We found phosphorylation of OPTN S342, PDCD4 S457, NUCKS1 T202, and MKI67 S1071 was upregulated in NR, whereas phosphorylation of TMEM45A S271, SH2D4A S124, OXR1 S202 and THRAP3 S406 was downregulated in NR. Except PDCD4 S457 which is phosphorylated by Akt and inhibits apoptosis, all p-sites are of yet unknown function. NUCKS1, encoded in 1q, is of unknown function but upregulated in many cancers and phosphorylated by CDK1 during mitosis. In conclusion, these p-sites might be potential biomarkers for patient stratification and play a functional role in responsiveness.

### Potential novel treatment options and drug targets

We searched for kinases driving resistance in our p-proteome data (Fig. S6E). AMPK activity was decreased in NR independent of treatment as determined by reduced phosphorylation of its substrates ACACA S80, RPTOR S722 and RAF S259. We confirmed significant downregulation of AMPK signaling in NR by performing PTM-SEA (Fig. 3E), which takes into account the actual p-site and not just the phosphorylated protein. Since AMPK inhibits proliferation, reduced AMPK activity in NR could contribute to aggressiveness and thus sorafenib resistance. AMPK activating drugs (AICAR) might therefore be of interest to counter sorafenib resistance. Interestingly, PTM-SEA also revealed decreased phosphorylation of p-sites in NR that are also targeted by the gamma-secretase inhibitor semagacestat and the HDAC inhibitor vorinostat. Gamma-secretase and HDACs are also part of the Notch signaling pathway and were also deregulated in NR, confirming our previous results for deregulated p-sites on Notch signaling components by PEA. Next, we searched a drugbank (Armstrong *et al*, 2020) for approved drugs that might antagonize sorafenib resistance (Fig. S6F). Drugs inhibiting proteins upregulated in NR included fedratinib (BRD4/FAK1), sunitinib/bosutinib (TNIK/FAK1) and olaparib (PARP1). Importantly, olaparib can overcome sorafenib resistance in HCC cells by suppressing the DNA damage repair signaling potentially through CHD1L (Yang *et al*, 2021). These results support further validation of drug combinations.

### Multi-omics integration segregates patients by therapy response and suggests increased dedifferentiation in nonresponder patients

To identify common molecular patterns of responsiveness, we performed data integration by multi-omics factor analysis (MOFA) (Argelaguet *et al*, 2018), which approximates the input data with a limited number of latent factors (LF). We observed a clear separation of NR and R along LF1 (Fig. 4A). Taken together, the difference between NR and R was manifested also by multi-omics integration, suggesting rather complex resistance mechanisms as opposed to single candidate genes or pathways.

**Fig. 4:**
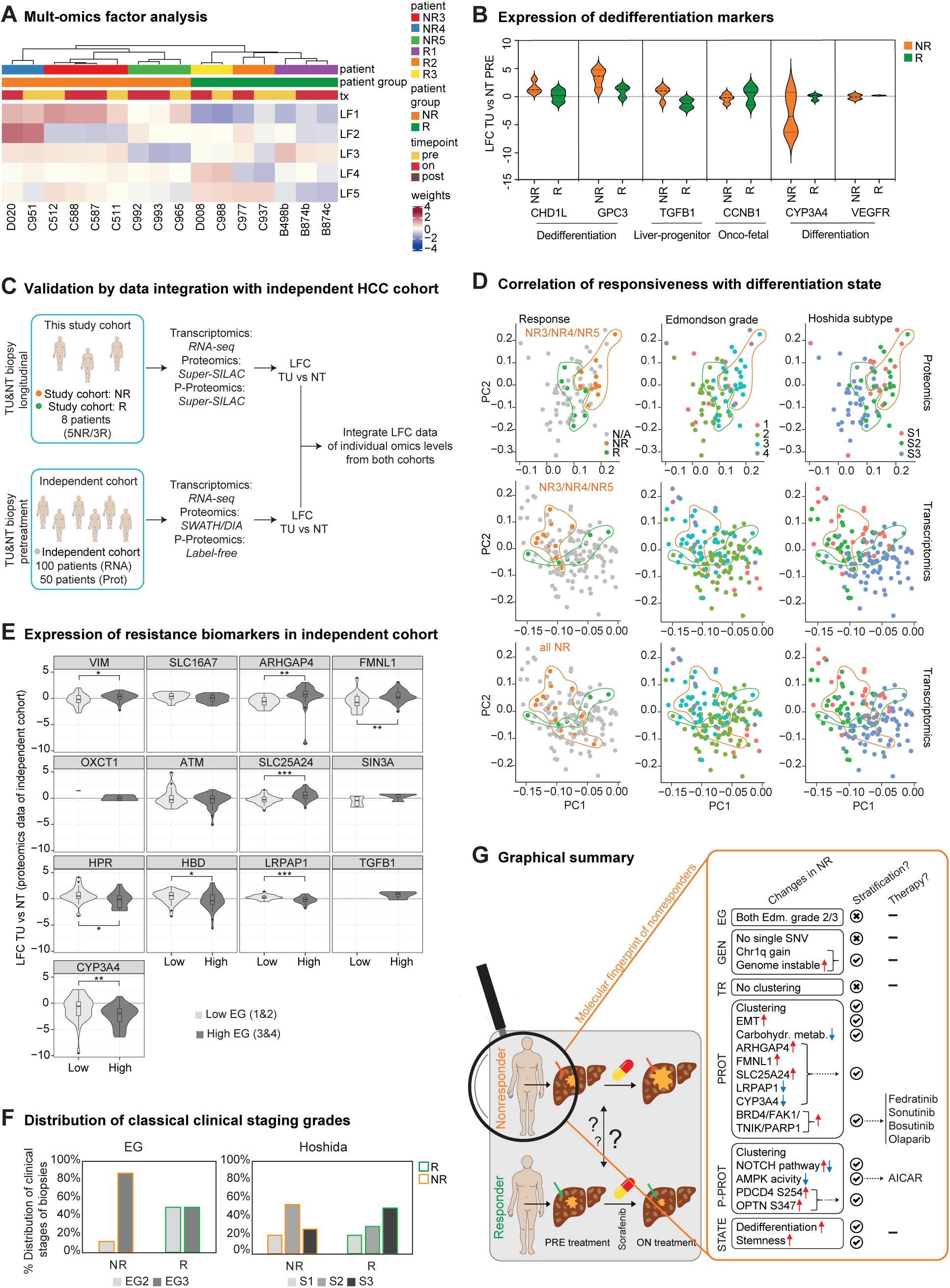
Multi-omics integration segregates patients by therapy response and suggests increased dedifferentiation in nonresponder patients. **(A)** HC of patient groups by the different latent factors (LFs) as obtained from MOFA analysis. Each dataset is composed of samples times features (e.g. proteins). MOFA explains dataset variance by weights corresponding to features and factors and by latent factors corresponding to factors and samples. The variance of the multi-omics dataset is described by red = positive weights, blue = negative weights. **(B)** Violin plot displaying pretreatment protein expression of selected hepatic de/differentiation markers in TU vs NT biopsies from NR and R patients. **(C)** Schematic representation of workflow to integrate this study cohort with the independent cohort of 100 (transcriptomics data)/50 ((p)proteomics data) HCC patients (Ng *et al*., 2021) for validation. **(D)** PCA of both integrated patient cohorts. Top row: proteomics data of all TU vs NT biopsies from main patients of this study (LFC) are labeled in green/red, middle row: transcriptomics data of all TU vs NT biopsies from main patients of this study (LFC) are labeled in green/red. bottom row: transcriptomics data from all TU vs NT biopsies from all patients of this study (LFC) are labeled in green/red. Left column: integrated LFC data of TU vs NT from both cohorts (independent cohort in grey), middle column: data points of both cohorts overlayed with colour of corresponding EG of biopsy, right column: data points of both cohorts overlayed with colour of corresponding Hoshida subgroup of biopsy. **(E)** Violon plot displaying selected potential novel protein biomarkers for responsiveness identified in study cohort. Their expression was analyzed in the proteome data from the independent cohort. Biopsies were grouped by EG low (grade 1 and 2) and high (grade 3 and 4). Statistical test between EG low and high group: Two-sample Wilcoxon test, ✶=p-value <0.05, ✶✶=p-value <0.01, ✶✶✶=p-value <0.001. CYP3A4 serves as positive control. **(F)** Distribution of clinical staging grades of biopsies (EG and Hoshida subclasses) between NR and R (all patient TU biopsies excluding metastases). **(G)** Graphical summary of this study.

To estimate tumor aggressiveness in NR, we analyzed protein expression of hepatic de-/differentiation markers (Liu *et al*, 2020) (Fig. 4B). Dedifferentiation markers CDH1L, GPC3 and TGFB1 were increased while differentiation markers CCNB1, CYP3A4 and VEGFR tended to be decreased in NR. Next, we investigated the possibility of increased dedifferentiation in NR by examining an independent cohort of pretreatment biopsies from HCC patients (transcriptome of 100 patients, (p)proteome of 50 patients) (Ng *et al*, 2021) (Fig. 4C). Indeed, NR tumors clustered as dedifferentiated tumors of higher Edmondson grades (EG) 3 and 4 and Hoshida subclasses 1 and 2 in the proteome and transcriptome of the independent cohort (Fig. 4D). Furthermore, seven out of twelve proteins strongly deregulated between NR and R were also significantly deregulated between EG low (1/2) and high (3/4) tumors of the independent cohort, e.g. ARHGAP4, FMNL1 and SLC25A24 (Fig. 4E). These proteins could be involved in hepatic dedifferentiation/tumorigenesis and serve as potential biomarkers for responsiveness.

Next, we quantified the distribution of clinical stages among our patients (Fig. 4F). 80% of NR tumors were EG3, while 50% of R tumors were EG3. On the Hoshida scale, 53% of NR tumors were S2 (aggressive type) while 27% of the R tumors were in this category. Thus, classical clinical staging alone does not unambiguously predict response to sorafenib.

## DISCUSSION

Here we describe the first longitudinal, proteogenomic study of HCC patients treated with the targeted kinase inhibitor sorafenib, with the aim of dissecting molecular differences between sorafenib responder and nonresponder patients (Fig. 4G).

We observed increased genomic instability, including a consistent 1q gain, in NR compared to R. Genomic instability is linked to chemoresistance and poor prognosis in other cancers, including HCC. 1q gain, a common event in HCC, is associated with reduced survival after therapy in multiple myeloma (Schmidt *et al*, 2019). Genomic instability or 1q gain could possibly stratify patients for sorafenib treatment, but may not be sufficient as our data suggest. It would possibly need to be combined with other molecular markers, for example those proposed in this study in a multi-level biomarker panel to test before treatment start Genomic instability might be a more general prognostic feature as well. Such hypothesis could be tested for example on the samples of the IMBRAVE −150 study.

We performed a multi-omics analysis on the study cohort. We did not find a single gene that confers resistance upon mutation but could correlate genomic instability with resistance. Furthermore, we found that proteomics and p-proteomics were superior to transcriptomics in segregating patients by therapy response. Therefore, transcriptomics or genomics alone (apart from CNA here) might not reveal functional differences between patient groups at sufficient depth. Using multi-omics in the clinics including (p-)-proteomics would therefore add important decisive information. In light of our data genomics and transcriptomics might even be dispensable for understanding the (lack of) response to sorafenib, since the proteome together with the p-proteome harbours enough critical information.

We discovered deregulated (non-canonical) NOTCH signaling and decreased AMP activity in NR. NOTCH signaling is implicated in HCC progression, poor prognosis, stemness signaling and EMT, but the underlying circuits remain poorly characterized (Villanueva *et al*, 2012). OXCT1, the rate limiting enzyme of ketolysis, was upregulated in NR. HCC upregulates OXCT1 and therefore ketolysis for progression under starvation. Enhanced ketolysis represses AMPK, thereby promoting tumor growth. Increased OXCT1 expression predicts higher patient mortality and upregulation of AMPK restores sorafenib sensitivity (Bort *et al*, 2019). NOTCH inhibitors, AMPK activators, fedratinib, sunitinib, bosutinib and rucaparib should be further explored to counter therapeutic sorafenib resistance.

We discovered molecular differences in NR, compared to R, potentially involved in therapeutic resistance. NR displayed high genomic instability, increased EMT, decreased metabolism, deregulated NOTCH signaling and reduced AMPK activity. Furthermore, NR expressed markers of poor prognosis (e.g., MCT2 and OXCT1), clustered with dedifferentiated tumors and expressed candidate resistance biomarkers correlating with dedifferentiation. Taken together, we hypothesize that responsiveness is linked to the differentiation status of the tumor. While genomic instability and EMT have been described to promote chemoresistance (also to sorafenib), we propose novel signaling cues potentially further promoting or regulating therapeutic resistance. Due to lack of mutations essential for resistance in our cohort, responsiveness could be driven by dynamic mutation-independent factors, e.g., stress responses or metabolic flux. Importantly, dedifferentiation was not evident from classical imaging-based clinical HCC staging. Since both NR and R contained EG2 and 3 pretreatment, such staging lacks molecular resolution and provides limited information for treatment decisions. Instead, staging via a molecular signature, e.g., our proposed candidate resistance biomarkers, may be more predictive. One could argue that increased dedifferentiation in NR is due to late HCC diagnosis and treatment start. However, sorafenib is given only to a clearly defined subgroup of late stage HCC patients (about 40%) (European Association For The Study Of The *et al*, 2012). Of note, we suggest potential intrinsic resistance biomarkers to stratify patients before treatment start and acquired resistance biomarkers to detect possible druggable targets deregulated only ontreatment. A large proportion of the differences between NR and R occured in an intrinsic manner, indicating that our proposed biomarkers are suited to detect patients intrinsically resistant to sorafenib.

Although our study is multi-layered and longitudinal, it has limitations due to its small cohort size of eight patients harbouring considerable intra- and inter-tumor heterogeneity. The small cohort size limits the power to detect statistically significant effects and bears the risk to overinterpret the observed differences between NR and R. To overcome this limitation, we integrated our study cohort with an independent cohort of HCC patients, thereby increasing both the scope and depth of our findings. Nevertheless, longitudinal tissue collection beyond the diagnostic HCC pretreatment biopsy is extremely rare, underlining the critical importance of our study as a resource for generating new testable hypotheses for the clinics.

In conclusion, we describe the first deep-scale multi-omic integration of a longitudinal HCC patient cohort. Our study demonstrates that proteomics together with p-proteomics can deepen the understanding of subtle yet important molecular differences between patients or patient groups. Several biomarkers discovered and quantified by proteomics are already applied in precision medicine, e.g. Ezrin used to predict lymph node metastasis in colorectal cancer (Su *et al*, 2021). The integration of (p)-proteomics in decision making processes at different stages in personalized medicine beyond genomics adds critical information with the potential to improve treatment outcome, as suggested by others (Doll *et al*, 2019).

## MATERIALS AND METHODS

### Study design and biopsy selection

Patients were recruited in the Clinic for Gastroenterology and Hepatology of the University Hospital Basel, Switzerland. They gave written informed consent to donate liver biopsy specimen for research. The study was approved by the Ethics committee of the Canton Basel, Switzerland, study protocol number EK 152/10. Biopsies were performed under ultrasound guidance using a coaxial needle technique allowing repetitive sampling from the same part of a focal lesion with a full-core biopsy instrument (BioPince®, Angitech, Stenlose, Denmark). HCC was diagnozed by histopathological analysis. Histopathologic grading was performed according to the Edmondson grading system (Edmondson & Steiner, 1954). Hematoxylin&eosin slides were reviewed to define presence or absence of cirrhosis, underlying liver disease etc. according to the guidelines by the World Health Organization. A detailed description about the clinical history of each patient, including which biopsies were taken at which timepoint and used for which analysis can be found in Figure S1A and B. The clinical parameters of each patient can be found in Table S1.

### DNA and RNA extraction

Genomic DNA and total RNA from tumor and adjacent liver parenchyma were extracted using the ZR-Duet DNA and RNA MiniPrep Plus kit (Zymo Research) following the manufacturer’s instructions. Prior to extraction, biopsies were crushed in liquid nitrogen to facilitate lysis. Total RNA of 19 patients without HCC was extracted using Trizol (Thermo Fisher Scientific) according to the manufacturer’s instructions. Extracted DNA was quantified using the Qubit Fluorometer (Invitrogen). Extracted RNA was quantified using NanoDrop 2000 spectrophotometer (Thermo Fisher Scientific), and RNA quality/integrity was assessed with an Agilent 2100 BioAnalyzer using RNA 6000 Nano Kit (Agilent Technologies).

### Whole-exome sequencing and data processing

Extracted DNA was sequenced using whole-exome sequencing. Whole-exome capture was performed using the SureSelectXT Clinical Research Exome (Agilent Technologies) or SureSelect Human All Exon V6+COSMIC (Agilent Technologies) platforms according to the manufacturer’s guidelines. Sequencing was performed on an Illumina HiSeq 2500 at the Genomics Facility Basel according to the manufacturer’s guidelines. Paired-end 101-bp reads were generated.

Sequence reads were aligned to the reference human genome GRCh37 using Burrows-Wheeler Aligner (BWA, v0.7.12) (Li & Durbin, 2009). Local realignment, duplicate removal and base quality adjustment were performed using the Genome Analysis Toolkit (GATK, v3.6) (McKenna *et al*, 2010) and Picard (http://broadinstitute.github.io/picard/). Somatic single nucleotide variants (SNVs) and small insertions and deletions (indels) were detected using MuTect (v1.1.4) (Cibulskis *et al*, 2013) and Strelka (v1.0.15) (Saunders *et al*, 2012). We filtered out SNVs and indels outside of the target regions: those with variant allelic fraction (VAF) of <1% and/or those supported by <3 reads. We excluded variants for which the tumor VAF was <5 times that of the paired non-tumor VAF. We further excluded variants identified in at least two of a panel of 123 non-tumor samples, including all non-tumor samples included in the current study, captured and sequenced using the same protocols using the artifact detection mode of MuTect2 implemented in GATK. To account for the presence of somatic mutations that may be present below the limit of sensitivity of somatic mutation callers, we used GATK UnifiedGenotyper to regenotype the positions of all unique mutations in all tumor samples from a given patient to define the presence of additional mutations supported by at least two reads. Mutations affecting hotspot residues (Chang *et al*, 2016) were annotated.

Allele-specific CNAs were identified using FACETS (v0.5.5) (Shen & Seshan, 2016), which performs a joint segmentation of the total and allelic copy ratio and infers allele-specific copy number states. Somatic mutations associated with the loss of the wild-type allele (i.e., loss of heterozygosity [LOH]) were identified as those for which the lesser (minor) copy number state at the locus was 0. All mutations on chromosome X in male patients were considered to be associated with LOH. Copy number states were inferred using two approaches (Piscuoglio *et al*, 2016). For the Log R ratio (LRR) approach, genes with copy number alterations were determined following the methods described in Curtis et al. (Curtis *et al*, 2012). In brief, the median Log2 ratio +2 standard deviations (SDs) or +6SDs was computed for the 50% (or 45% or 40%, see below) of the central positions ordered by their Log2 ratios to define copy number gains and amplifications, respectively, and the median Log2 ratio −2.5SDs or −7SDs was computed for the 50% (or 45% or 40%) of the central positions to detect copy number losses and homozygous deletions, respectively. To account for the differences in tumor cell content, ploidy and noise between samples, the proportion of central positions, ordered by their Log2 ratios, was determined based on the median absolute difference between the raw Log2 ratio and the segmented Log2 ratio. For samples in which the median absolute difference was ≤0.2, >0.2 and ≤0.3, central positions of >0.3, 50%, 45% and 40% were adopted, respectively. For the allele-specific CNA (ASCNA) approach, the FACETS copy number states following the expectation-maximization procedure were used. Copy number states were collapsed to the gene level based on the median values to coding gene resolution based on all coding genes retrieved from the Ensembl (release GRCh37.p13). Genes with ASCNA total copy number 0 were considered homozygously deleted.

### Analysis of copy number events

Gene-specific CN calls were obtained by taking the consensus of the gene-specific LRR and ASCNA calls: a gene was considered lost if both LRR and ASCNA calls agreed on a loss, and gained if both calls agreed on a gain or amplification. Samples were excluded due to poor quality when more than 100 genes were called homozygously deleted. Plotting was done with the R package ComplexHeatmap.

### Survival curves

Genomic and clinical data for the selected sorafenib-treated HCC dataset (Harding *et al*., 2019) were downloaded from cbioportal (https://www.cbioportal.org) in October 2021. Only sorafenib-treated patients were taken into account. Differences in patient survival (OS and PFS) were assessed using the Kaplan-Meier method and analyzed using the log-rank test as previously described (Kancherla *et al*, 2018). All tests were two-sided. P < 0.05 were considered statistically significant. Statistical analyses were performed with Prism 9.2.0 (GraphPad Software).

### RNA-sequencing and data processing

RNA-seq library prep was performed with 200 ng total RNA using the TruSeq Stranded Total RNA Library Prep Kit with Ribo-Zero Gold (Illumina) according to manufacturer’s specifications. Single-end 126-bp sequencing was performed on an Illumina HiSeq 2500 using v4 SBS chemistry at the Genomics Facility Basel according to the manufacturer’s guidelines. Primary data analysis was performed with the Illumina RTA version 1.18.66.3. Sequence reads were aligned simultaneously to the human reference genome GRCh37, HBV strain ayw genome (NC 003977.2), and HCV genotype 1 genome (NC 004102.1) by STAR (Dobin *et al*, 2013) using the two-pass approach.

Transcript quantification was performed using RSEM (Li & Dewey, 2011). Gene-level expected counts were upper-quartile-normalized to 1000. For downstream analysis, we computed the log_2_-fold-changes of normalized RSEM gene counts between each sample and the median of 15 normal livers.

### Preparation of super-SILAC mix for quantitative (p)proteomics

A super-SILAC (Geiger *et al*, 2010) mix was prepared from HCC cell lines: Huh7 of high and low passage, HepG2, Huh6 and Hep3B, kindly provided by Prof. Ralf Bartenschlager (University of Heidelberg, Germany). For labeling, cells were grown in DMEM with 10% dialyzed FCS (MW cut-off 10 kDa), 100 IU/ml penicillin, 100 μg/ml of streptomycin, 2 mM L-glutamine, 1x non-essential amino acids without proline, containing heavy (R10K8, ^13^C- and ^15^N-labeled) or light amino acids (R0K0). After 8-10 passages, label incorporation was tested by LC/MS/MS. Upon 95-100% labeling, cells were expanded to prepare large amounts of heavy super-SILAC mix. At 80-90% confluence, cells were harvested and lysed. Protein concentration was determined by Bradford assay, and cell extracts from each cell line were mixed at a 1:1:1:1:1 ratio. Aliquots were snap frozen in liquid nitrogen and stored at −80°C.

### Protein extraction and digestion

Fresh biopsies were immediately snap-frozen in liquid nitrogen. Sample preparation for mass spectrometry was performed as described before (Dazert *et al*, 2016). In brief, for protein extraction, each frozen biopsy was crushed in an in-house constructed metal mortar cooled on dry ice into a fine powder (cryogenic grinding) and transferred into a cooled 1.5 ml tube containing 150 - 400 μl lysis buffer (50 mM Tris-HCl pH 8.0, 8M urea, 150 mM NaCl, 1 mM PMSF, Complete Mini Protease inhibitor (Roche) and phosphatase inhibitor Phos-Stop (Roche)). The biopsy lysate was vortexed vigorously for 5 min, rotated for 1h at 4°C and sonicated twice for 1 min in a UP200St µltrasonic processor (Hielscher). After lysis samples were centrifuged for 10 min at 10,000 x g in a table top centrifuge at 15°C and supernatant was taken. The pellet was then further homogenized. To this end, additional 50µl of lysis buffer were added to the pellet and samples were homogenized with a hand-held pellet pestle (Kimble) twice for 30 sec on ice. Samples were again centrifuged for 10 min at 10,000 x g in a table top centrifuge at 15°C. Supernatants were pooled with the first cleared lysate. Protein concentration was measured with a Bradford assay. Heavy super-SILAC mix (Geiger *et al*., 2010) was added in a 1:1 ratio to the light protein lysate. Next, proteins were reduced with 10 mM DTT for 1h at 37 °C and alkylated with 50 mM iodoacetamide for 30 min at RT in the dark, both with gentle shaking. Urea concentration was lowered to 4 M with 50 mM Tris-HCl, pH 8.0. Lysates were digested with two rounds of endoproteinase LysC (Wako) at a 1:100 enzyme-to-protein ratio at 37°C for two hours. Next, the urea concentration was lowered to 1 M. Lysates were digested with two rounds of trypsin (Promega): 1:50 ratio overnight and 1:100 ratio for 2 hrs at 37°C. A second trypsin aliquot (1:100 ratio) was added for 2.5 hrs at 37°C. Digestion was stopped with TFA to a final concentration of 0.5%. Digests were centrifuged for 2 min at 1,500 g and desalted on a C18 SepPak cartridge (50mg column for up to 2.5mg peptide load capacity) (Waters) (Dephoure & Gygi, 2011). Peptide concentration was estimated at 280nm on a nanodrop spectrophotometer (IMPLEN) and peptides were dried in the SpeedVac.

### Strong cation exchange chromatography (SCX)

SCX fractionation was done according to (Dephoure & Gygi, 2011) with modifications. The dried peptides were resuspended in 1.5 ml of SCX buffer A (5 mM KH_2_PO_4_, pH 2.65, 30% AcCN), briefly sonicated and centrifuged at 10,000 rpm. The HiTrap SP cartridge (GE Healthcare) was equilibrated three times with 1ml of SCX buffer A, then washed three times with 1ml of SCX buffer B (5 mM KH2PO4, pH 2.65, 30% AcCN containing 500 mM KCl) and re-equilibrated three times with 1ml of SCX buffer A. Peptides were applied onto the column and washed three times with 1 ml SCX buffer A. Flow through and washes were collected separately. The bound peptides were stepwise desorbed with 1 ml each of SCX buffer A containing 50 mM, 100 mM, 150 mM, 250 mM, 350 mM, and 500 mM KCl (optionally 10 mM, 25mM) and the fractions were collected individually. The OD of each fraction was measured at 280 nm. Fractions were dried in a SpeedVac and desalted on C18 columns (The Nest Group) of varying size adjusted to the peptide content. 20% of each fraction, but maximum 30µg, were separated for LC/MS/MS analysis as the proteome.

### P-peptide enrichment

P-peptide enrichment was performed with titanium-dioxide (TiO_2_) coupled beads (GL Sciences Inc.) as described before (Kettenbach & Gerber, 2011). The p-peptide pools were desalted on MicroSpin columns (see above) and dried in the SpeedVac without heating.

### LC/MS/MS analysis

The first training patient (NR1) was measured on an Orbitrap Classic instrument (Thermo Scientific) with the LC/MS settings described previously (Dazert *et al*., 2016), the second training patient (NR2) was measured on a QExactive Plus instrument (Thermo Scientific) with the same LC/MS settings as described before (Dazert *et al*., 2016). The remaining patients (NR3, NR4, NR5 and R1, R2, R3) were also analyzed on a QExactive Plus instrument (Thermo Scientific) with the following optimized LC/MS settings. The dried (p)peptides were dissolved in 20-30 μl of 0.1% formic acid and injected into the LC/MS. Proteomes and p-proteomes were analyzed by capillary liquid chromatography tandem MS (LC/MS/MS) using a homemade separating column (0.075 mm x 38 cm for proteome and 0.075 mm x 20 cm for p-proteome) packed with Reprosil C18 reverse-phase material (2.4 μm particle size, Dr. Maisch, Ammerbuch-Entringen, Germany). For the proteome, the column was heated with an in-house made column oven to 55°C. The peptides were analyzed on a Thermo Scientific QExactive Plus instrument coupled to an Easy nLC 1000 capillary pump (Thermo Scientific). The column was connected on-line and the solvents used for peptide separation were 0.1% formic acid in water (solvent A) and 0.1% formic acid/80% AcCN in water (solvent B). The proteome was measured with a metal T-cross and the p-proteome with a rubber T-cross to avoid p-peptide loss. 2 μl of resuspended (p)peptides were injected to a maximum pressure of 500 bar for the proteome and 350 bar for the p-proteome. For proteome measurements, a linear gradient from 0 to 40% solvent B in solvent A in 190 min was delivered with the nano pump at a flow rate of 200 nl/min. After 190 min solvent B was increased to 95% in five minutes. For p-proteome measurements, a two-step gradient from 0 to 15% solvent B in solvent A in 55 min followed by an increase to 40% solvent B in solvent A in 40 min was delivered with the nano pump at a flow rate of 200 nl/min. After 95 min solvent B was increased to 95% in 25 min. The eluting peptides were ionized at 2.5 kV. The mass spectrometer was operated in data-dependent mode. The precursor scan was done in the Orbitrap set to 70,000 resolution, and the MS2 scan was done at a resolution of 17,500 for the proteome and at 35,000 for the p-proteome. For the proteome, both maximum injection times (precursor scan and fragment ion scan) were set to 120 ms, for the p-proteome the first was set to 60 ms and the second to 120 ms. A top twenty method was run for both proteome and p-proteome.

### Protein identification and data processing

The LC/MS/MS data were analyzed with MaxQuant, version 1.5.1.2 (Cox & Mann, 2008) and searched against the human SwissProt database (29.10.2013). The default settings of MaxQuant were used, except the following deviations: Multiplicity was set to 2 and Arg10 and Lys8 were set as labels; pSTY was set as variable modification; Main search peptide tolerance was set to 10ppm; Match-between-run and Re-quantify option was enabled; PSM FDR, Protein FDR and Site decoy fraction was set to 0.02; Protein quantification minimal ratio count was set to 1. All samples belonging to the same patient were analyzed together, but with an individual analysis for the proteome and p-proteome. An experimental design template was compiled, where all runs belonging to the same biopsy were pooled as one parameter group and all runs belonging to the same fraction were labeled as same fraction. The protein and pSTY datasets were exported into a FileMaker Pro 12 data bank. Normalized ratios H/L were taken for all analyses. Only p-sites with a localization probability of >50% were taken into account (class I/II/III sites). All ratios were log_2_-transformed and median subtracted.

The proteome and p-proteome of a total of 48 biopsies from 8 patients were measured. All biopsies from one patient were measured together (proteome and p-proteome separate) to ensure similar LC/MS/MS conditions for all fractions. The number of IDs detected in NR1 and NR2 were much lower compared to the rest of the patients, due to technical/instrumental improvements for the latter. Therefore, NR1 and NR2 proteome and p-proteome data were excluded from downstream analysis, but are still available as “training patients”, e.g. to validate a hypothesis. All biopsies from NR3, NR4, NR5, R1, R2 and R3 (“main patients”) passed the QC criteria, namely 1) comparable TIC counts and 2) comparable chromatogram shapes of the different SCX fractions and 3) clear subgrouping with the same tissue type biopsies on the PCA. Since we measured all biopsies, we collected data for multiple ontreatment time points from one patient. We decided to use only one pre- and one ontreatment timepoint per patient, but allowed multiple biopsies from the same timepoint for PEA and candidate search using (p)proteomics data.

### Identification of Top deregulated proteins and p-sites between NR and R

For the proteome, we extracted all proteins that were measured in at least 70% of the biopsies per patient group per time point and then performed a bidirectional Students T-test between both groups. Next, we filtered the proteins with a LFC>0.75, then sorted the list by p-value and chose the Top 500 proteins. In order to perform pathway enrichment, we used a background list of proteins that were quantified at least in one pair of TU vs matched NT per patient group. The Top 50 list was extracted from the Top 500 list by filtering for the best p-values.

For the p-proteome, we first filtered for the p-site with the best q-value per protein to condense the list to one p-site per protein. Next, we followed the same procedure as for the proteome. However, due to the lower recovery in the p-proteome, we extracted a Top 300 list for the analysis including all timepoints together (overall) and a Top 200 list for the individual pre- and ontreatment analysis. In order to perform pathway enrichment analysis, we used a background list of p-sites that were quantified in at least one biopsy per patient group per timepoint. The Top 50 list was extracted from the Top 500 list by filtering for the best p-values.

### Pathway enrichment analysis of (p)proteome data

Pathway enrichment analysis (PEA) for the Top proteins and p-sites deregulated between nonresponders and responder patient data was performed with WebGestalt (http://bioinfo.vanderbilt.edu/webgestalt/ (Zhang *et al*, 2005). Settings for WebGestalt were default except the following: An over-representation analysis (ORA) was performed using KEGG and Reactome pathway database and the Top 15 hits were shown as a result. For the proteome, the background list consisted of all protein which were quantified in at least one TU and NT biopsy per group (nonresponder and responder group) and for the p-proteome, the background list consisted of those p-sites quantified in at least one biopsy per group (nonresponder and responder group). Important pathways from WebGestalt analysis were chosen to visualize pathway changes (e.g. EMT, NOTCH signaling). Involved proteins/p-sites were placed as “Omics tiles” in these pathways by manual building up the pathways from literature. The architecture of the MAPK pathway was adapted from (Matallanas *et al*, 2011; McCubrey *et al*, 2007; Mendoza *et al*, 2011), of the mTOR pathway was adapted from (Mendoza *et al*., 2011; Shimobayashi & Hall, 2014) and of the NOTCH pathway was adapted from (Borggrefe & Oswald, 2009; Bray, 2006, 2016; Kopan & Ilagan, 2009).

### Statistical analysis of (p)proteome data

For statistical data analysis, hierarchical cluster analysis, principal component analysis and Volcano plots the Perseus program, version 1.6.14 (Cox & Mann, 2012), was used. For the volcano plots, a bidirectional Students t-test was performed, adjusting S0 to 1, number of randomizations to 250 and FDR to 2% for the proteome and to 5% for the p-proteome. For the heatmaps, an ANOVA multi-sample t-test was performed, adjusting S0 to 1, number of randomizations to 250 and FDR to 2% for the proteome and to 5% for the p-proteome. Z-scoring was performed without grouping. For unsupervised hierarchical clustering, the distance was calculated by Spearman, the linkage set to average and the maximal numbers of clusters to 300. Z-score ranges of the corresponding heatmaps can be found in Data File S2.

### Multi-omics database for manual curation of candidates

Transcriptomics, proteomics and p-proteomics data were compiled into a Filemaker (Filemaker Pro Advanced, Claris) database. To find proteins and p-sites involved in responsiveness to therapy, this database was manually curated for proteins or p-sites rated as differentially expressed between TU and matched NT by three hit criteria (1) LFC >0.7 in TU vs matched NT and significant in bidirectional Students T-test (p-value>0.02 in proteome and >0.05 in p-proteome) or (2) LFC >1 in TU vs matched NT but not necessarily significant or (3) «Black-and-white» protein or p-site, which means that a protein or p-site was quantified in all three triplicate measurements of the TU or NT biopsy and quantified in none of three triplicate runs in the matched TU or NT biopsy. Proteins were curated for affiliation to one of six different molecular patterns of regulation: NR down R up (extreme pattern), NR down R unchanged, NR unchanged R down, NR up R down (extreme pattern), NR up R unchanged, NR unchanged R up. Each candidate needed to be quantified in >2 TU biopsies per group. For the extreme patterns, a candidate needed to be changed adversely in >50% of the TU biopsies in both groups (nonresponder and responder group) and in the rest of the patterns it needed to be changed in >70% of the TU biopsies of only one group and unchanged in the other group.

### Choice of potential protein or p-site biomarkers for responsiveness

From the FMP search (search done in Filemaker database for manual curation of candidates) only the members of the most extreme pattern groups (see paragraph before) were chosen as potential biomarkers. The FMP search for biomarkers was only performed for the pre- and ontreatment timepoint, but not the combined data.

From the LFC search (search done by choosing the Top50 candidates from the protein or p-site list which is based on Log2 fold change of TU/NT and p-value) only members of the most extreme pattern groups (see hierarchical tree for proteins or p-sites on the left site of the corresponding heatmap) were chosen. The LFC search for biomarkers was only performed for the pre- and ontreatment timepoint, but not the combined data. From the extreme pattern groups, only those candidates with a z-score range <-1.5 to >1.5 were chosen.

### Kinase substrate enrichment analysis (KSEA)

To perform KSEA we first merged the two major publicly available databases for documented Kinase– Substrate Relations (KSRs) from SIGNOR (https://signor.uniroma2.it) and PSP (https://www.phosphosite.org). Since the database obtained from the SIGNOR platform provides more detailed information about each KSR we used the SIGNOR database as base and added all KSRs that are present in the PSP database but absent in the SIGNOR database. After integrating the combined KSR information into our Omics database we exported the obtained upstream Kinase information as semicolon separated categories for further analysis in Perseus (1.6.5.0). After filtering p-sites that do have KSR information we could perform Anova analysis of the remaining 840 p-sites based on the LFC between nonresponders and responders, taking into account all timepoints together. For the Anova analysis we used an FDR of 10% and an S0 value of 0.1 as threshold and the number of randomizations was set to 250. This resulted in 78 p-sites that are significantly upregulated and 67 p-sites that are significantly downregulated in NR vs R. With these two groups and the background list of 840 p-sites we performed the enrichment analysis for upstream kinases and used the resulting p-values to rank the associated kinases to estimate the likeliness of the respective kinase to be activated or deactivated.

### PTM-SEA analysis

TU vs NT phosphoproteomic profiles of responder and nonresponder patient groups were aggregated separately by averaging the log_2_ fold changes on the p-site-level. The two scores and the associated p-sites were then used with ssGSEA (version 2.0, Git revision b3d035e) to conduct independent PTM-SEA (version 1.9.0) analyses. The Fig. was generated using ggplot2, labelling gene sets (FDR<0.25) in each quadrant, where opposite responder *vs* nonresponder enrichment directionality is depicted. Significant (FDR<0.05) gene sets are highlighted in red. Opacity indicates significance (FDR).

### Drug database analysis

To find drugs that target proteins strongly deregulated in NR, we consulted the drug database www.guidetopharmacology.com (Armstrong *et al*., 2020). We analyzed in total 2970 described drug-target relations. From each treatment timepoint (pre, on and overall), we generated two lists of the Top 50 deregulated proteins, one from the LFC search and one from the FMP search (only pre and on) (see above). From both lists, we chose only the proteins upregulated in NR compared to R (approx. 50% of each list). We then searched for drugs that fulfill two criteria: 1) the drug is described to target one or more of the upregulated proteins and 2) the drug is already approved.

### Multi-omics factor analysis (MOFA)

For the integrative multi-omics analysis using MOFA, we considered all data modalities, limiting the DNA to CN events, due to the high levels of heterogeneity in the SNV data. CN events (gains and losses) found in at least two patients were kept, and summarised in a binary matrix, where 1 denoted presence of the event. Genes displaying the same pattern of CN alteration across the 29 samples were condensed into a single CN event, leading to 147 events (85 gains, 62 losses).

RSEM estimates of gene expression were loaded into DESeq2 where a variance-stabilizing transformation (VST) was applied. The VST was blind to the experimental design so as not to bias the values with information about the response group for the downstream unsupervised analysis. Only protein-coding genes and miRNAs were kept. Proteins and p-sites were measured in triplicate. Only features with two valid measurements were considered and averaged to produce their summarised value. Summarised values, ratios of protein to spike-in, were transformed with a binary logarithm and each sample was centred by subtracting its mean.

LFCs were computed for RNA and (p)proteome to address patient-specific effects (such as sex or a varying number of samples) that we would otherwise be unable to handle due to the limited number of available samples. LFCs were computed for each tumour sample against its matched non-tumour sample. For every data modality except DNA, the 500 most variable features across patients were selected by their largest squared coefficient of variation.

MOFA2 (Argelaguet *et al*, 2020; Argelaguet *et al*., 2018) was run with default parameters except that scale_views was set to true because of the different variance of the RNA data compared to the (p)proteome, num_factors was set to 8, maxiter was set to 10,000 and convergence_mode was set to “slow”.

### Comparison of the HCC study cohort and the independent HCC cohort

#### Analysis of transcriptomic profiles

All tumor and nontumor transcriptomic RNA-seq profiles of the this study cohort and the independent cohort ((Ng *et al*., 2021) were reanalyzed using Kallisto (Bray *et al*, 2016) (version 0.46.2) and the human Ensembl (version 103) transcriptome index with parameters “-b 100 --single -l 150 -s 25”. Differential gene expression analysis was conducted using tximport (Soneson *et al*, 2015) (version 1.16.1) and edgeR (Robinson *et al*, 2010) (version 3.30.3), pairwise comparing single nontumor and tumor samples to obtain fold-changes.

#### Analysis of p-proteomic profiles

The tumor and nontumor p-proteomic profiles of the independent cohort (Ng *et al*., 2021) were reprocessed using MSFragger (Kong *et al*, 2017) (version 3.1.1), Philosopher (da Veiga Leprevost *et al*, 2020) (version 3.3.11) and IonQuant (version 1.4.6) (Yu *et al*, 2020) against the canonical human UniProtKB/Swiss-Prot database (obtained 2021-02-10) with appended decoys and contaminants. Quantile normalization was applied for the three batches separately and non-p-peptides were excluded for all further analysis. Only runs covering more than 1,000 p-sites were considered for further analysis. The log_2_-transformed peptide precursor intensities were averaged across the three replicates. Peptide intensities were median centered for each sample and then the log_2_-fold-change was computed on p-peptide-level between tumor and corresponding nontumor samples.

#### Comparison of transcriptomic samples

Missing LFC values were imputed with 0, and then principal component analysis and visualization of the first two components was conducted using “prcomp” from the R-packages “stats” (version 4.0.3) and “ggplot2” (version 3.3.3), respectively.

#### Comparison of proteomic and p-proteomic samples

To improve comparability between the two studies, which were measured by different proteomic platforms, the analysis results were restricted to the same p-sites or proteins, respectively. To account for different peptide or protein detection rates within the three batches of the classification study, detected peptides or proteins were subsampled to ensure identical detectability distributions for each peptide or protein. Subsampling was then also conducted between the two datasets. Missing LFC values were imputed with 0, and then principal component analysis and visualization of the first two components was conducted using “prcomp” from the R-packages “stats” (version 4.0.3) and “ggplot2” (version 3.3.3), respectively.

## Data availability

1. The genomics (CNA and SNV) and transcriptomics data of the longitudinal study cohort have been deposited in the European Genome-phenome archive (EGA) with the accession IDs EGAS00001005661 (WES) and EGAS00001005662 (RNA).
2. The proteomics and phosphoproteomics data of the longitudinal study cohort have been deposited to the ProteomeXchange Consortium via the PRIDE (Perez-Riverol *et al*, 2019) partner repository with the dataset identifier PXD028593.
3. The proteomics and phosphoproteomics data of the independent cohort (data partially published in Ng *et al*., 2021) have been deposited to the ProteomeXchange Consortium via the PRIDE (Perez-Riverol *et al*, 2019) partner repository with the dataset identifier PXD034593 and PXD035238.

## Acknowledgments

We thank the patients for donating their samples for research.

## Funding

European Research Council Synergia grant 609883 MERiC (M.H., N.B., M.N.H)

Swiss National Science Foundation (M.N.H)

Goldschmidt-Jacobson-Stiftung (E.D.)

## Author contributions

E.D., N.B., M.H. and M.N.H. designed experiments. T.B., S.W. and M.H. collected clinical samples. T.B., C.K.Y.N, S.W. and T.B. generated patient database. L.T. performed pathology evaluation of liver samples. S.W., T.B. and S.K. performed genomics and transcriptomics analysis. C.K.Y.N. performed mutation calling of genomics data and quantification of transcriptomics data, E.D. performed proteomics and phosphoproteomics ((p)proteomics) analysis, E.D. and M.C. analyzed (p)proteomics data. M.C. assembled and curated omics database. E.D., M.C., F.M. and G.R. performed statistical analysis of proteogenomics data. E.D. interpreted proteogenomics and integrated multi-omics data. F.M. and N.B. performed MOFA and SNV/CNA analysis. S.P. performed survival analysis. G.R. performed PTM-SEA and integration of study cohort and independent cohort. E.D. and M.N.H. wrote the manuscript.

## Competing interests

Authors declare that they have no competing interests

## SUPPLEMENTARY MATERIAL

**Supplemental Fig. S1.**
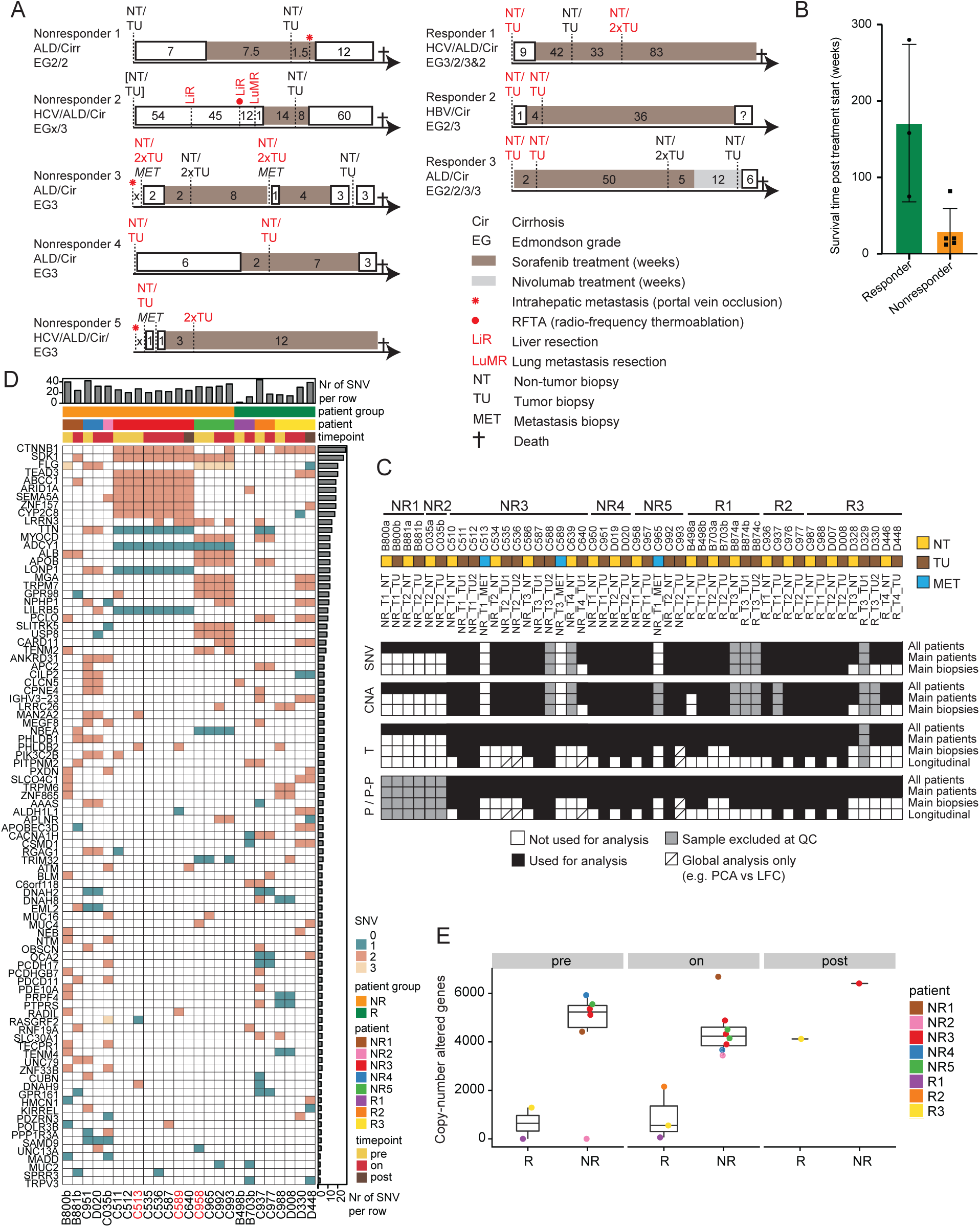
Patient histories, sample usage and SNV/CNA analysis of eight sorafenib-treated HCC patients. (A) Patient histories of eight sorafenib-treated HCC patients. Biopsies coloured in red were used for transcriptomics, proteomics and p-proteomics analyses (main biopsies). (B) Survival time post treatment start of responder and nonresponder patients used in this study cohort. See Table S1. (C) Sample usage of patient biopsies in the different omics analyses in this study. (D) Synonymous and non-synonymous SNVs are shown which appeared in the TU biopsies of a minimum of two patients. Patient columns were ordered by therapy response, gene rows were ordered by row sums from top to bottom. Column header displays column sums; row sides display genes and row sums. Portal vein metastases biopsies are labeled in red. Synonymous SNVs are shown in green, non-synonymous SNVs in dark orange, if a gene is affected by both types, it is shown in pale yellow. (E) Number of CNAs per biopsy and time point. Biopsies from same patient are labeled in same colour.

**Supplemental Fig. S2:**
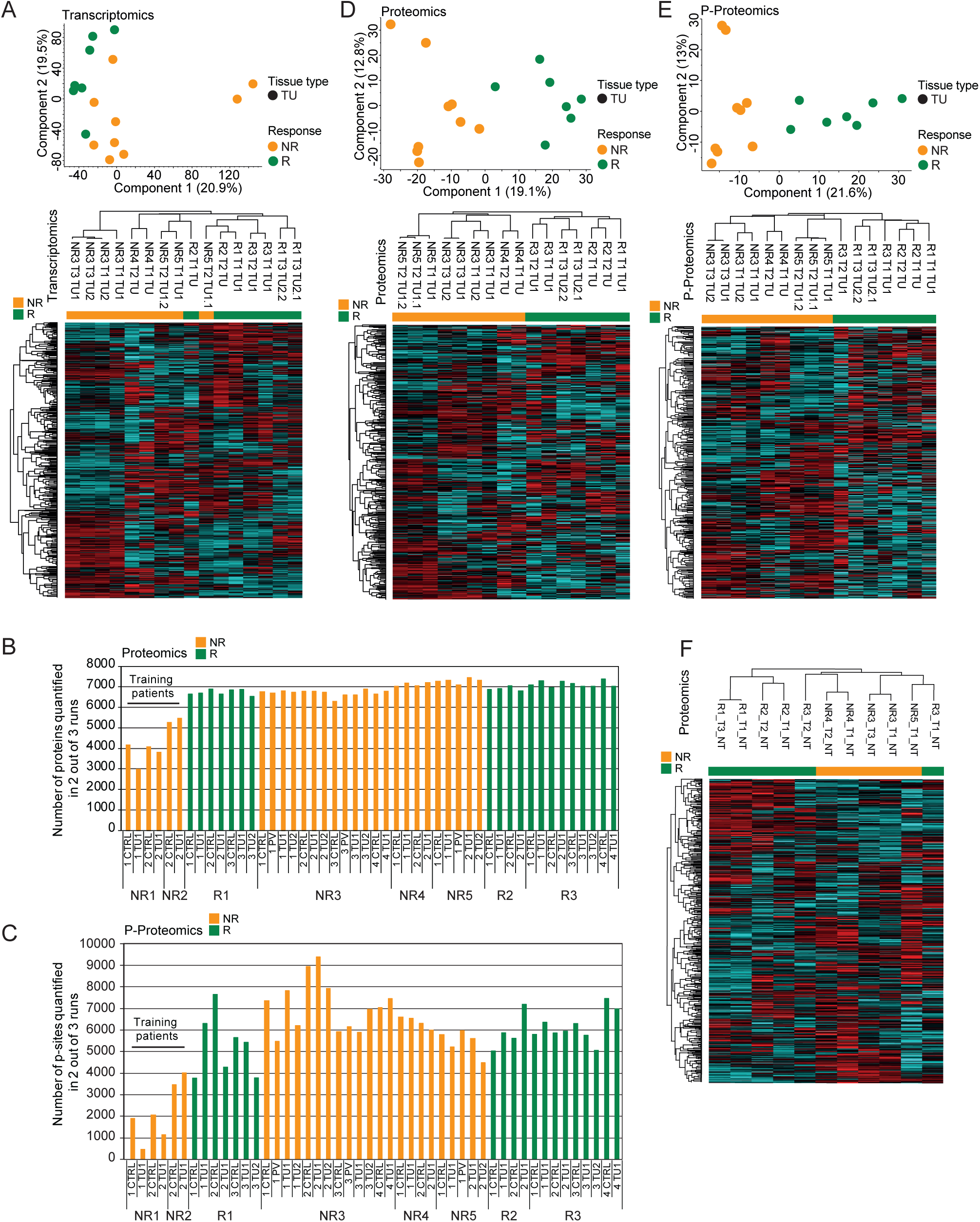
Patients clustered by therapy response, based on the proteome and p-proteome but not the transcriptome. (A) Analyses of transcriptomics data, namely mean ratios from TU biopsies of main patients. Upper panel: PCA, bottom panel: HC. Shown are mean ratios of each biopsy i.e., mean of triplicate measurements of the normalized ratio H/L. Z-score ranges shown Table S2. (B, C) Statistics of proteomics (B) and p-proteomics (C) measurements by LC/MSMS. Patients are ordered by chronological measurement. Each bar corresponds to the mean of triplicate measurements. Nonresponder 1 and 2 are “training patients”. These biopsies were measured on different mass spectrometers, resulting in lower protein ID recovery (see method section). (D, E) Analyses of proteomics (D) and p-proteomics (E) data, namely mean ratios from TU biopsies of main patients. Upper panel: PCA, bottom panel: HC. Shown are mean ratios of each biopsy i.e., mean of triplicate measurements of the normalized ratio H/L. Z-score ranges shown Table S2. (F) HC of proteomics data, namely mean ratios from NT biopsies of main patients. Shown are mean ratios of each biopsy i.e., mean of triplicate measurements of the normalized ratio H/L. Z-score ranges shown Table S2.

**Supplemental Fig. S3:**
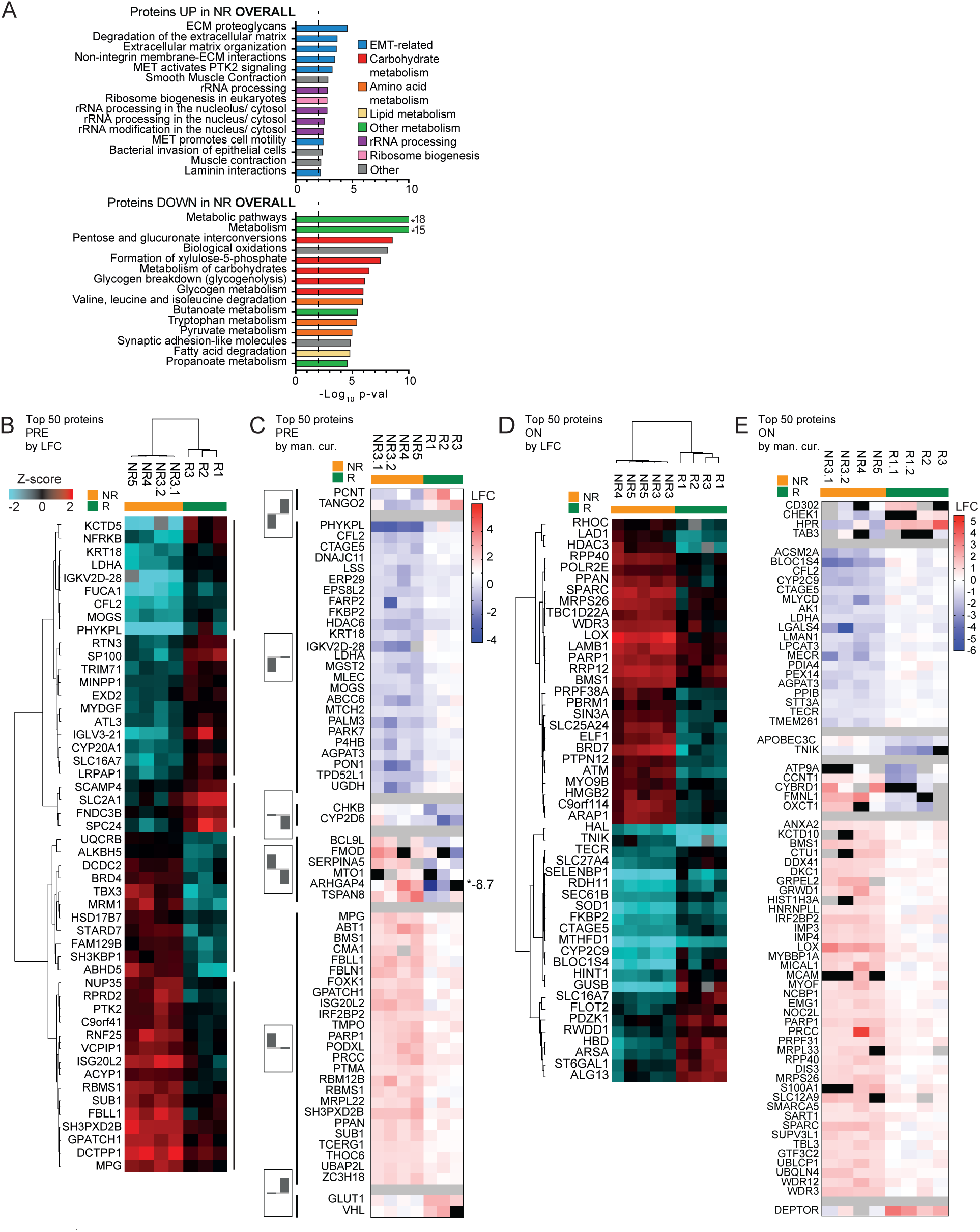
Pathways and proteins deregulated on proteome level. (A) PEA of protein changes deregulated overall (all timepoints together) between NR and R based on p-value and LFC of TU vs NT (Top 500). Top panel: upregulated proteins, Bottom panel: downregulated proteins. Pathway annotations belonging to the same cellular process were labeled with the same colour. (B, D) HC of the Top 50 deregulated proteins differentially expressed pretreatment (B) and ontreatment (D) between NR and R based on p-value and LFC TU vs NT. Z-score ranges shown Table S2. (C, E) Manual curation of proteins rated as differentially expressed pretreatment (C) and ontreatment (E) between NR and R by three possible hit criteria. (1) LFC>0.7 in TU vs NT and significant (p-value<0.02) (2) LFC>1 in TU vs NT but not necessarily signficant (3) Black-and-white protein (B&W). Proteins were curated for affiliation to one of six different molecular patterns (see left side of (C)): NR down R up, NR down R unchanged, NR unchanged R down, NR up R down, NR up R unchanged, NR unchanged R up. See methods section for details.

**Supplemental Fig. S4:**
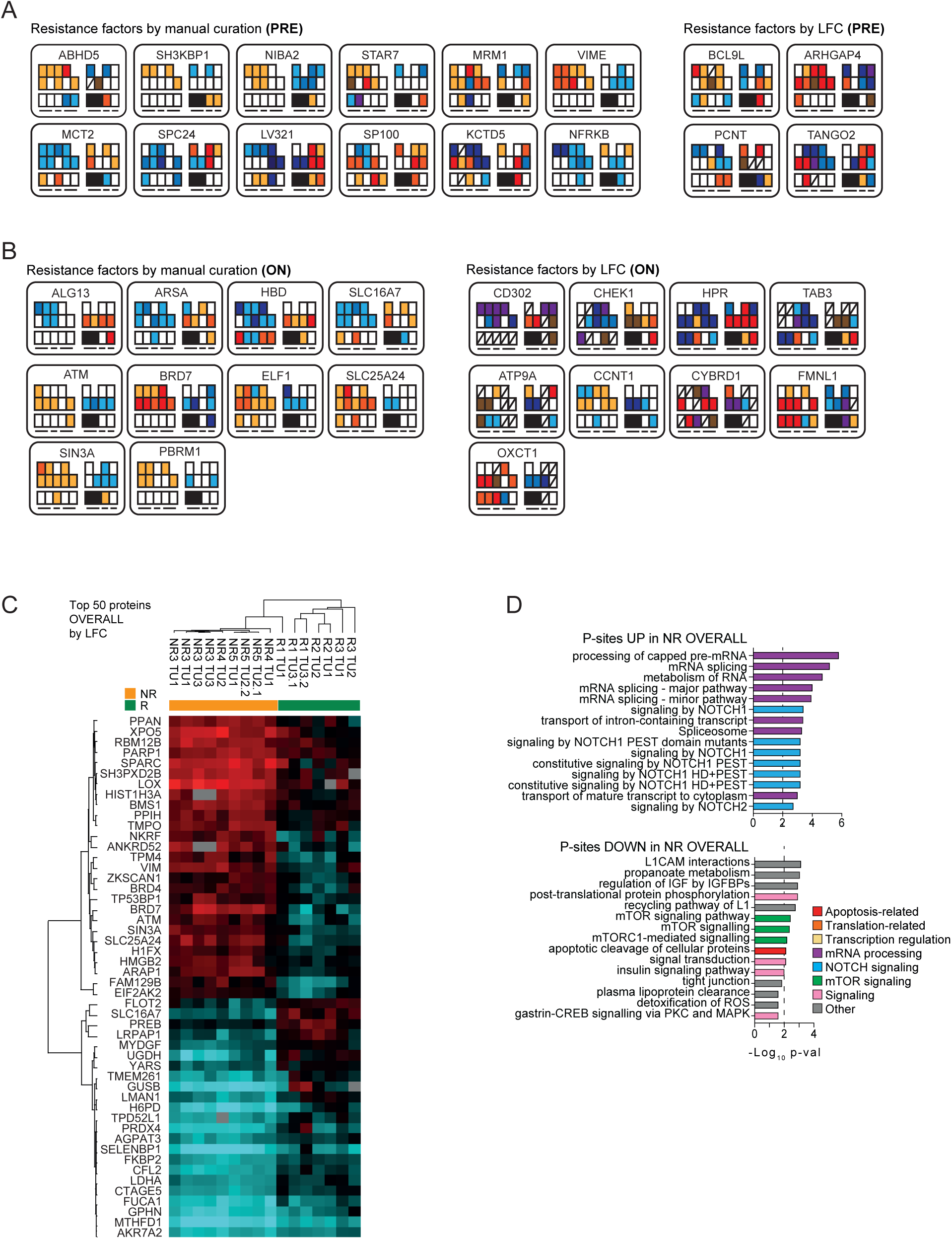
Pathways, proteins and p-sites deregulated on proteome and p-proteome level. (A, B) Omics tiles of potential protein biomarkers for responsiveness. Shown are strongest deregulated proteins between NR and R. Pretreatment candidates (A) extracted from Fig. S3B and S3C and ontreatment candidates (B) extracted from Fig. S3D and S3E. Left side: candidates identified by manual curation, right side: candidates identified by LFC. (C) HC of the Top 50 proteins differentially expressed overall (all timepoints together) between NR and R patient groups by p-value and LFC. Heatmap shows Z-scored LFC of TU vs matched NT biopsy. Z-score ranges shown Table S2. (D) PEA of proteins that carry the Top 300 deregulated p-sites differentially phosphorylated overall (all timepoints together) between NR and R patient groups by p-value and LFC. Top panel: upregulated p-sites, middle panel: downregulated p-sites. Pathway annotations belonging to the same cellular process were labeled with the same colour. Dashed line indicates significance threshold.

**Supplemental Fig. S5:**
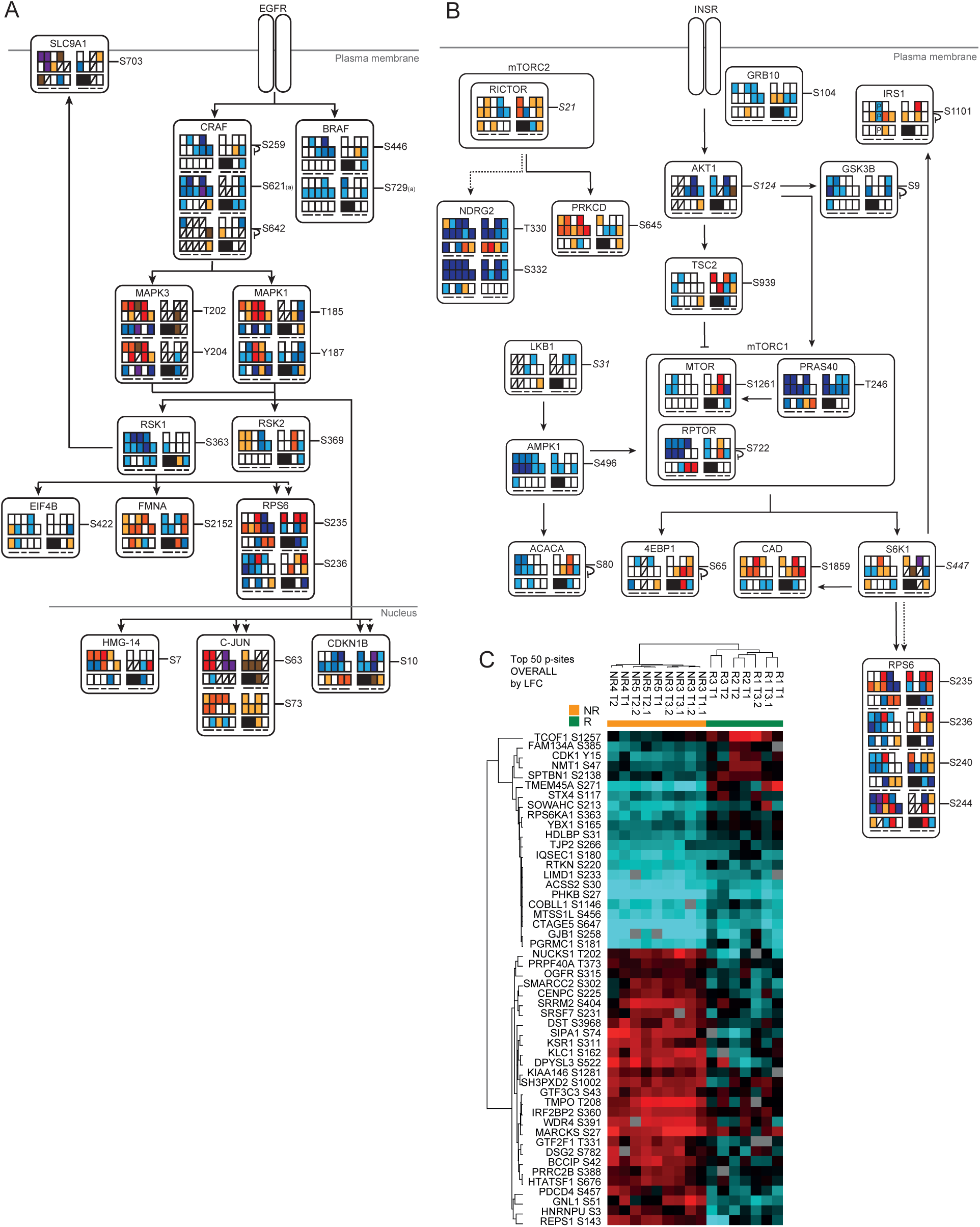
Pathways and p-sites deregulated on p-proteome level. (A) Omics tiles for p-sites involved in MAPK pathway (adapted from (Matallanas *et al*., 2011; McCubrey *et al*., 2007; Mendoza *et al*., 2011)). Fig. displays p-sites that were quantified in this study. Straight arrow indicates phosphorylation of target, small round arrow with blank end indicates inhibitory auto-phosphorylation. (B) Omics tiles of p-sites belonging to the mTOR pathway (adapted from (Mendoza *et al*., 2011; Shimobayashi & Hall, 2014)). Straight arrow indicates phosphorylation of target, small round arrow with blank end indicates inhibitory auto-phosphorylation. Dashed line indicates 1) that phosphorylation of this p-site can also be performed by additional kinase or 2) that another kinase in between phosphorylates this site. (C) HC of the Top 50 deregulated p-sites differentially phosphorylated overall (over all timepoints together) between NR and R patient groups based on p-value and LFC TU vs NT. Heatmap shows Z-scored LFC of TU vs matched NT biopsy. Z-score ranges shown Table S2.

**Supplemental Fig. S6:**
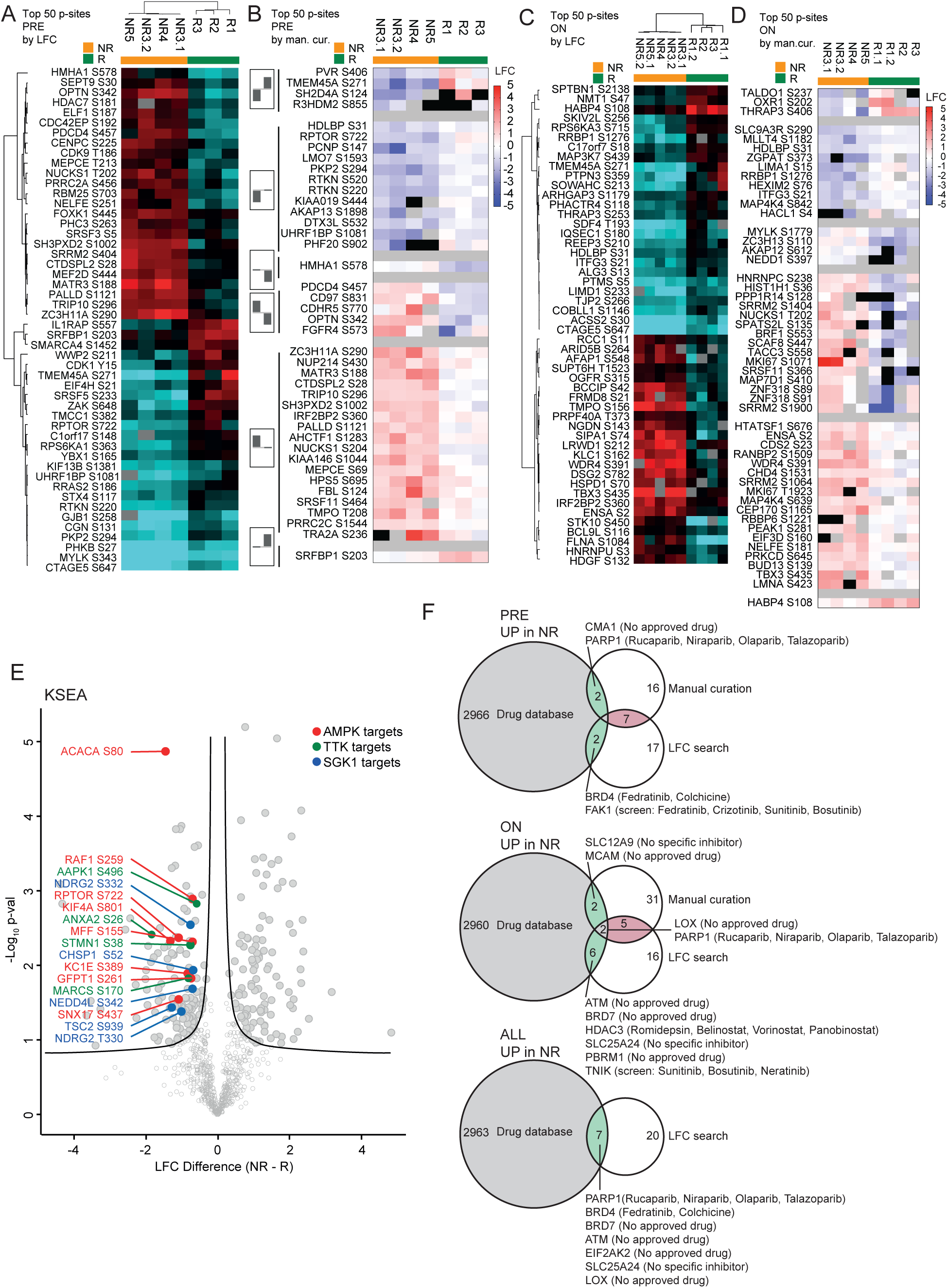
P-sites and kinases deregulated on p-proteome level and potential combinatorial drugs to counter sorafenib resistance. (A, C) HC of the Top 50 deregulated p-sites differentially phosphorylated pretreatment (A) and ontreatment (C) between NR and R based on p-value and LFC TU vs NT. Z-score ranges shown in Table S2. (B, D) Manual curation of p-sites rated as differentially phosphorylated pretreatment (B) and ontreatment (D) between NR and R by three possible hit criteria. (1) LFC>0.7 in TU vs NT and significant (p-value<0.02) (2) LFC>1 in TU vs NT but not necessarily signficant (3) Black-and-white protein (B&W). Proteins were curated for affiliation to one of six different molecular patterns (see left side of (B)): NR down R up, NR down R unchanged, NR unchanged R down, NR up R down, NR up R unchanged, NR unchanged R up. See methods section for details. (E) Kinase-substrate-enrichment analysis (KSEA) of p-sites deregulated between NR and R overall (taking into account all timepoints together). Highlighted are p-sites with decreased phosphorylation in NR tumors compared to R which are targets of the kinases AMPK, TTK or SGK1. (F) Overlap of known drug-target relations from the drug database Guide to Pharmacology (https://www.guidetopharmacology.org) <u>with the Top 50 proteins deregulated between NR and R.</u> Only proteins upregulated in NR were taken into account. Upper panel: pretreatment. Middle panel: ontreatment. Lower panel: overall (taking into account all timepoints together). Manual curation = manual curation of FMP database with three hit criteria (see above and Methods section), LFC search = Top 50 list of proteins sorted by p-value and LFC of TU vs NT (see above and methods section).

**Supplementary Table S1:**
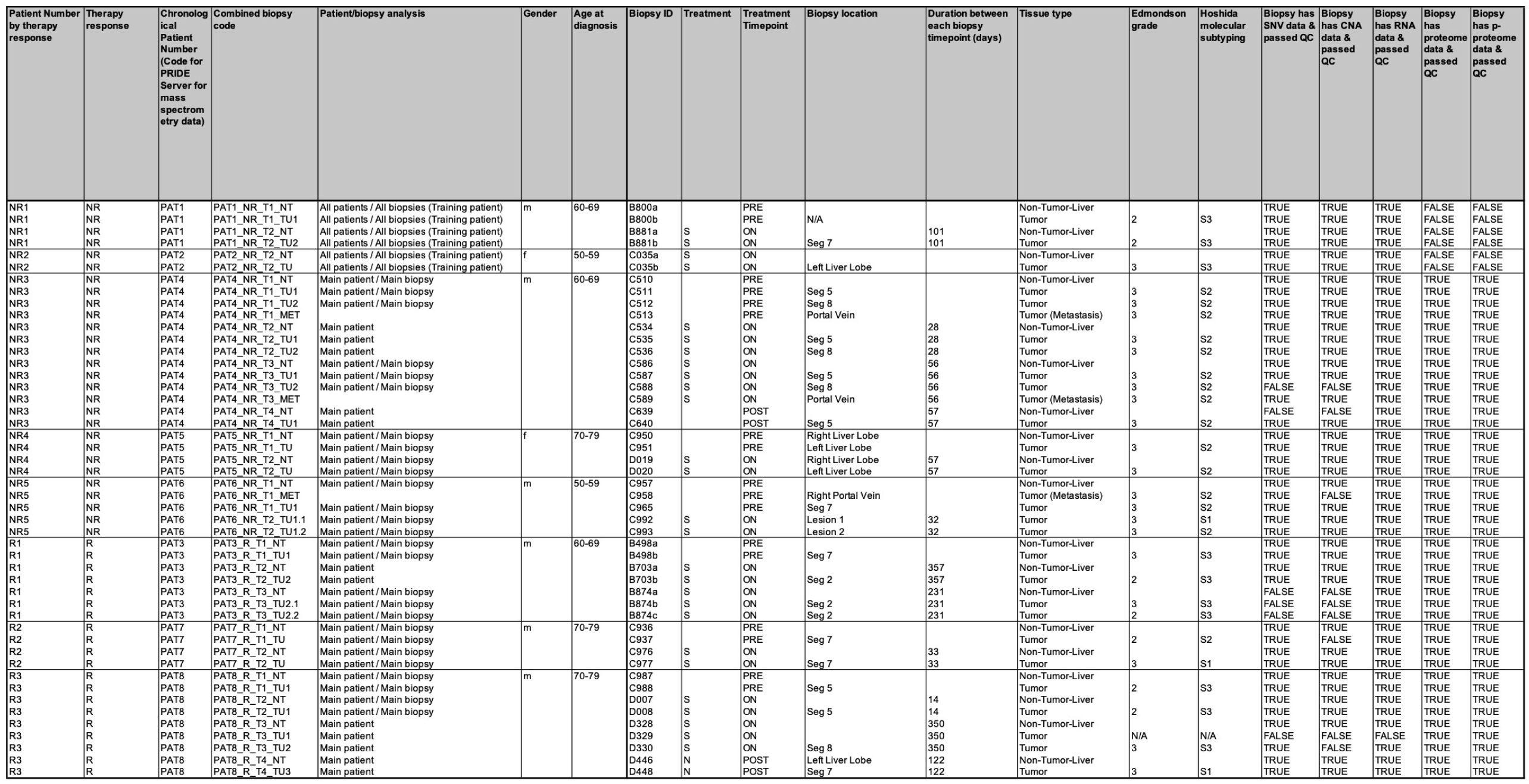
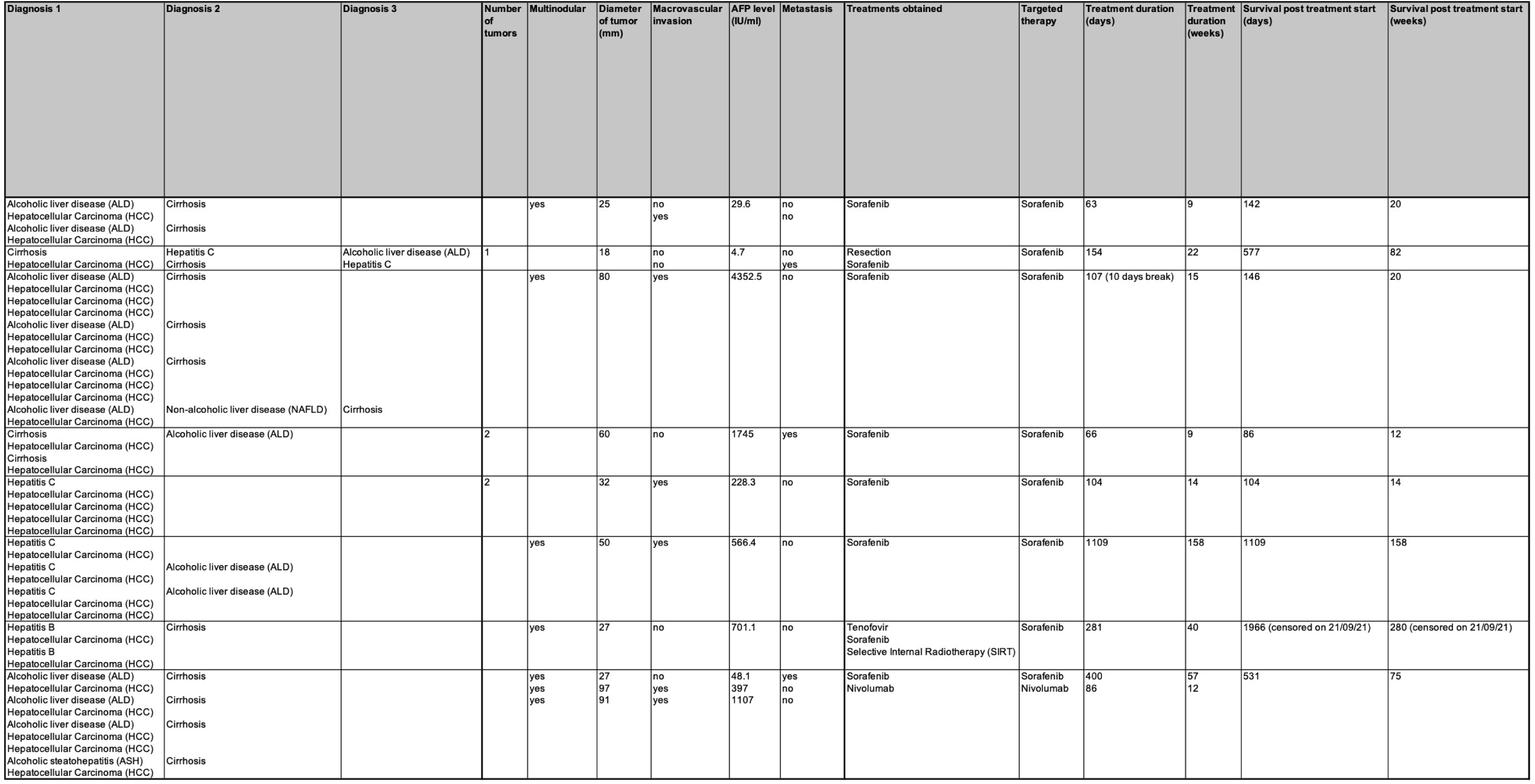
Clinical parameters of eight sorafenib-treated HCC patients. NR = Nonresponder patient R = Responder patient NT = nontumor biopsy TU = tumor biopsy T1 = pretreatment biopsy T2 = first ontreatment biopsy T3 = second ontreatment biopsy T4 = biopsy after therapy TU1,2,3 = Tumor biopsies from different nodules of same patient TU1.1, T1.2 = Two tumor biopsies from same nodule MET = metastasis biopsy (portal vein) S = under sorafenib therapy

**Supplementary Data file S1: Z-score ranges of hierarchical clusterings shown in this study**

See XLS sheet

**Supplementary Data file S2: Top 500 deregulated proteins between NR and R (PRE, ON and OVERALL)**

See XLS sheet

**Supplementary Data file S3: Top 200/300 p-sites deregulated between NR and R (PRE, ON and OVERALL)**

See XLS sheet

## Notes

### Competing Interest Statement

The authors have declared no competing interest.

### Funding Statement

This study was funded by European Research Council Synergia grant 609883 MERiC (M.H., N.B., M.N.H). Other funding: Swiss National Science Foundation (M.N.H), Goldschmidt-Jacobson-Stiftung (E.D.)

### Author Declarations

The Ethics committee of the Canton Basel, Switzerland gave ethical approval for this work

